# Sex-specific survival bias and interaction modeling in coronary artery disease risk prediction

**DOI:** 10.1101/2021.06.23.21259247

**Authors:** Ida Surakka, Brooke N Wolford, Scott C Ritchie, Whitney E Hornsby, Nadia R. Sutton, Maiken Elvenstad Gabrielsen, Anne Heidi Skogholt, Laurent Thomas, Michael Inouye, Kristian Hveem, Cristen J Willer

## Abstract

**Background:** The 10-year Atherosclerotic Cardiovascular Disease (ASCVD) risk score is the standard approach to predict risk of incident cardiovascular events and recently, addition of CAD polygenic scores (PGS_CAD_) have been evaluated. Although age and sex strongly predict the risk of CAD, their interaction with genetic risk prediction has not been systematically examined.

**Objectives:** This study performed an in-depth evaluation of age and sex effects in genetic CAD risk prediction.

**Methods:** The population-based Norwegian HUNT2 cohort of 51,036 individuals was used as the primary dataset. Findings were replicated in the UK Biobank (372,410 individuals). Models for 10-year CAD risk were fitted using Cox proportional hazards and Harrell’s concordance index, sensitivity, and specificity were compared.

**Results:** Inclusion of age and sex interactions of PGS_CAD_ to the prediction models increased C-index and sensitivity likely countering the observed survival bias in the baseline. The sensitivity for females was lower than males in all models including genetic information. The two-step approach identified a total of 82.6% of incident CAD cases (74.1% by ASCVD risk score and an additional 8.5% by the PGS_CAD_ interaction model).

**Conclusion:** These findings highlight the importance and complexity of genetic risk in predicting CAD. There is a need for modeling age and sex-interactions terms with polygenic scores to optimize detection of individuals at high-risk, those who warrant preventive interventions. Sex-specific studies are needed to understand and estimate CAD risk with genetic information.

**CONDENSED ABSTRACT:** This study used two large population-based longitudinal datasets to evaluate genetic prediction of CAD including age and sex interactions. The model fit and sensitivity of the prediction models increased when including age and sex interaction of PGS_CAD_ to the prediction models likely countering the observed survival bias in the baseline. The sensitivity for females was lower than for males in all models including genetic information. Our results highlight the importance and complexity of genetic risk and suggest including age and sex interactions with polygenic scores to identify more high-risk individuals for preventive interventions.

## INTRODUCTION

Coronary artery disease (CAD) is a complex disease influenced by risk factors including hypertension, hyperlipidemia, diabetes, tobacco use, age, and genetics, which leads to high morbidity and mortality(1). The American College of Cardiology / American Heart Association(2) recommends the Pooled Cohort Equation (PCE, or the 10-year Atherosclerotic Cardiovascular Disease [ASCVD] risk score) to estimate an individual’s risk using several demographic and cardiovascular disease risk factors. Other models include Systematic COronary Risk Evaluation (SCORE)(3), QRISK(4), Framingham risk score(5), and NORRISK(6). The predictive capacity of these models is moderate (C-index is between 0.6-0.8), depending on characteristics of the external validation dataset such as age and statin use(7-10).

There is significant additive value of integrating genome-wide genetic data to enhance risk prediction using polygenic scores (PGS) (7-9(10)). Additionally, individuals with a PGS in the highest 8% of score distribution have a risk of CAD comparable to having monogenic familial hypercholesterolemia (3-fold increased risk) (10). To date, investigators have shown that adding PGS(11-17) to standard risk prediction algorithms enhances the power of the model to predict CAD, consistent with the estimated contribution of genetic factors responsible for 40-50% of CAD risk(18).

The most predictive components of CAD prediction models are age and sex(2). The interplay of these two factors with the other traditional risk factors has been evaluated extensively in epidemiologic studies(2-6). However, careful consideration of age and sex interactions has not been systematically applied to genetic risk prediction. We used a longitudinal population-based dataset of 51,036 samples from Norway and performed Cox proportional hazards models to explore whether age and sex impact CAD genetic risk prediction. The objective was to identify whether CAD genetic risk scores’ performance in the prediction of incident CAD depends on patient’s age and sex.

## MATERIALS AND METHODS

### Study Cohort

The Trøndelag Health Study(19) (HUNT) has collected samples during three different time periods: HUNT1 (1984-1986), HUNT2 (1995-1997), and HUNT3 (2006-2008). Participation in HUNT is based on informed consent, and the study has been approved by the Data Inspectorate and the Regional Ethics Committee for Medical Research in Norway (REK: 2014/144). HUNT1 was excluded because lipid panels were not available for this cohort while HUNT3 was excluded given a median follow-up time of less than 10 years. HUNT2 was the primary dataset (N= 80,658) for this analysis.

Individuals with complete baseline information at the time of study enrollment, including cohort characteristics **(Supplemental Table 1)**, hospital registry data, and genotype data available were included. The definition of CAD can be found from **Supplemental Methods**. Individuals with prevalent CAD at baseline were excluded. The final dataset consisted of 51,036 individuals between the ages of 19 to 99 years (median follow-up, 21.2 years). To estimate 10-year risk of CAD, the longitudinal data analyses were restricted to the first 10 years of follow-up. All but one (non-CAD related death during follow-up, censored in the analyses) of non-cases had a full 10 years of follow-up. HUNT2 genotyping was performed using Illumina Human CoreExome v1.1 array with 70,000 additional custom content beads and imputed from a combined imputation panel including HRC and 2,202 low-pass HUNT genomes using an approach described previously(20).

### Polygenic Risk Score Calculation

The CAD polygenic score (PGS_CAD_) used here is based on metaGRS weights from Inouye et al.(12). MetaGRS shows best performance metrics of the CAD scores in the PGS Catalog from where the weights were downloaded (https://www.pgscatalog.org). The PGS_CAD_ was calculated as the weighted sum of effect alleles using the reported weights (w_m_)

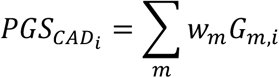

where G_m,i_ is the dosage of effect alleles of individual i for marker m. The resulting raw score follows a Gaussian distribution **(Supplemental Figure 1A)** and was adjusted with the first 10 genetic principal components (**Supplemental Figure 1B**). The adjusted score was further inverse-normal transformed for the analyses to have the hazard ratios (HR) on the standard deviation (SD)-scale unless stated otherwise. The inverse normal transformation was performed in males and females separately for the sex-specific models. The PGS_CAD_ does not include sex chromosome variants.

### ASCVD risk score

ASCVD risk was calculated using weights provided by American Heart Association Taskforce guidelines(21). In models where ASCVD risk was evaluated with the PGS_CAD_, the ASCVD risk was fitted into a Cox Proportional Hazards model as a continuous variable. The ASCVD values used in the Cox models range between (0,1) instead of percentages. As previously reported(22,23), ASCVD risk tends to overestimate the CAD risk for individuals in the highest risk groups. Individuals with a predicted risk ≥ 7.5% for ASCVD were considered medium to high risk. This is also the threshold at which lipid lowering therapy is clinically-indicated in the United States (24). Using this threshold, the miscalibration observed in the ASCVD risk for those with high risk estimates should not have had a noticeable effect on the reclassification metrics **(Supplemental Figure 2 A-B)**.

### Replication cohort

The United Kingdom (UK) Biobank dataset was used to replicate our findings. A full description of the dataset has been previously described(25). For this study, the dataset was restricted to individuals with European ancestry as the PGS_CAD_ weights were from an association study of European ancestry only. Individuals with prevalent CAD events and individuals on lipid lowering medication were excluded from the analysis. Samples that were used to train the metaGRS in the original publication(12) were excluded to avoid possible bias. The final dataset had 372,410 individuals, of which 17,569 had an incident CAD event or CAD-related death over the 10.9-year follow-up. The CAD definition used in UK Biobank can be found in the **Supplemental Methods**. The UK Biobank cohort descriptive statistics can be found in **Supplemental Table 2**. All statistical models in UK Biobank were adjusted for baseline assessment center to account for possible geographical biases and most recent nation of abode to account for differences in follow-up time available for hospitals in England, Wales, and Scotland.

### Statistical Methods

We used three main model types: linear models, Cox Proportional Hazards models for the combined data, and Cox Proportional Hazards models stratified by sex. The linear models were used to examine the non-time-dependent correlation structures between the variables of interest at baseline, whereas the Cox Proportional Hazards models were applied to examine the time-dependent predictiveness of the variables over the 10-year follow-up. **Supplemental Table 3** summarizes the Cox models, including the models with combined data (models C1-C4) and the models stratified by sex (models S1-S3).

To compare the different models to each other and their possible utility in clinical practice, we used the concordance index (Harrell’s C-index, referred to as C-index throughout the study), sensitivity, and specificity. C-index is a model fit statistic for survival models that is a generalization of the receiver operating characteristic curve that also handles censored data. In practice, the higher the C-index, the better the estimated risk is in concordance with the observed risk (i.e., individuals with high predicted risk are incident cases, and those with low risk are non-cases). However, sensitivity and specificity are dependent on the assigned risk threshold (7.5% in our study). Sensitivity is the proportion of cases assigned into the high-risk category while specificity is the proportion of non-cases in the low-risk category.

All statistical analyses were performed in R version 3.6.3 (https://cran.r-project.org) with downloadable libraries survival (https://CRAN.R-project.org/package=survival) and PredictABEL (https://CRAN.R-project.org/package=PredictABEL). All HUNT2 participants were ascertained between 1995-1997, and therefore, it was not necessary to account for the possible baseline time-period effects. As such, the Cox Proportional Hazards models were fit with follow-up time as the time-scale using R function coxph().

## RESULTS

### CAD Polygenic Score and Correlations with Age and Sex

We initially tested whether the genome-wide polygenic score for CAD estimated using metaGRS weights(12) (hereafter called the PGS_CAD_) was associated with sex or age in HUNT2 before examining how to optimally account for age and sex in CAD risk-stratification model. In principle, one would expect PGSs for various diseases to show equivalent distributions among males and females, provided age, sex, and ancestry are corrected in the underlying summary statistics, and sex-chromosomes are excluded from the evaluation. However, the relationship between the PGS, sex, and age could be impacted by ascertainment bias, undetected population stratification, or survival bias by genotype.

By considering baseline data only, we observed significant associations between enrollment age and PGS_CAD_ and between sex and PGS_CAD_ (**Supplemental Table 4A-B**). These results suggest that there is non-random selection in the cohort related to PGS_CAD_, possibly ascertainment bias or survival effects. Based on the results from these two models, males at baseline had an average 0.06 SD-units lower PGS_CAD_ than females, and the PGS_CAD_ was 0.003 SD-units lower per year of age. The association of age with PGS_CAD_ was significant for both males and females (**Supplemental Table 4C-D**), although the effect for males was marginally higher (PGS_CAD_ 0.0032 SD-units lower per year for males, 0.0025 for females) **(Figure 1A)**. However, upon adding the interaction term to the linear regression model, age*sex term was not significant (P-value = 0.212). The sex-PGS_CAD_ association was partly age dependent as the effect of sex on PGS_CAD_ becomes non-significant (P-value = 0.404) when the interaction term is added (**Supplemental Table 4E**). The age and sex associations were confirmed by testing the models in the UK Biobank. In UK Biobank the interaction term age*sex on the PGS_CAD_ was statistically significant (P-value = 1.0e-8; **Supplemental Table 5A-C, Supplemental Figure 3A)**. The trends were reduced when including the prevalent cases (together with the baseline statin users in the UK Biobank) in the baseline analysis in both datasets, (N=1,455 in HUNT2, N=84,292 in UK Biobank; **Figure 1B, Supplemental Table 6A-B, Supplemental Figure 3B**). The slight gradual decrease in mean PGS_CAD_ by age, particularly in men, could be due to lower survival of older males with high PGS_CAD,_ who are probably absent from the cohort at baseline (and some of whom were excluded from analyses due to prevalent or earlier-onset CAD).

**Figure 1.**
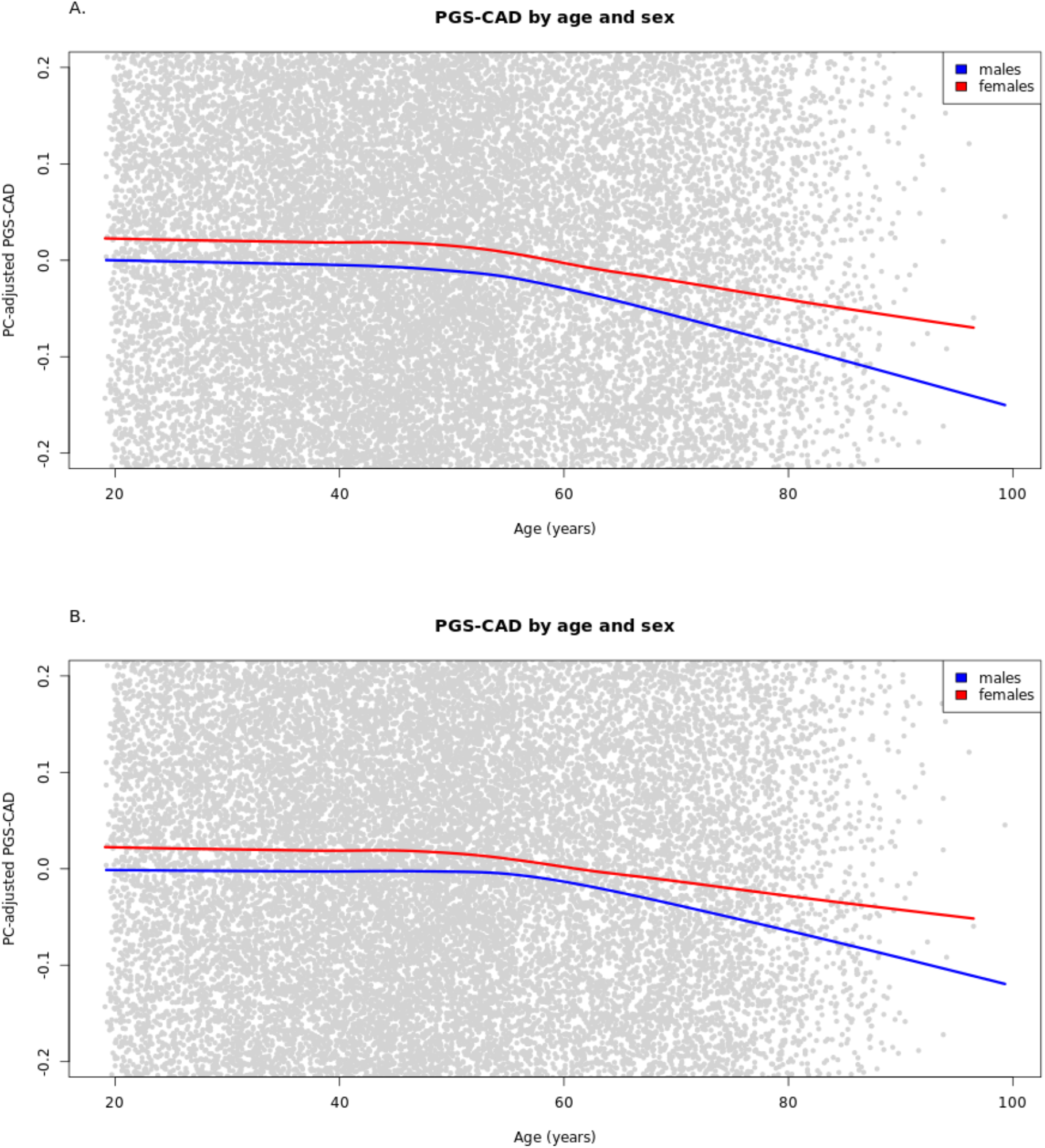
Raw PGS_CAD_ by age and sex in the cohort baseline. Illustration of the selection bias in the cohort baseline using lowess curves for PGS_CAD_ by age for males and females separately. The plot has been zoomed in relative to the y-axis to better show the trends. Panel A shows the trends in the analysis dataset used in the Cox models (prevalent cases excluded) and panel B when prevalent cases are included.

### CAD Polygenic Score and Age and Sex Interaction Models

We tested PGS_CAD_ performance while explicitly modeling age, sex, and a comprehensive set of interaction terms to counter the survival bias observed at baseline. We used an additive PGS_CAD_ model (model C1, **Supplemental Table 8**) as the comparison model to test the effect of added interaction effects to the model performance (**Supplemental Table 9**). To fully capture any potential interactions, we examined a model including all interaction terms of age, sex, and PGS_CAD_ to test for the possible age-dependent (Age*PGS_CAD_ term) and sex-dependent (Sex*PGS_CAD_ term) behavior of the PGS_CAD_, and age-effects of the PGS_CAD_ predictive performance that may differ between males and females (Age*Sex*PGS_CAD_ term) (model C2).

The sensitivity (78.4%) increased in the full interaction model (model C2, **Supplemental Table 10**) compared to the model with additive genetic effects only (model C1) (sensitivity 77.0%, **Table 1**) whereas the C-index did not show significant increase (model C2 C-index 0.839 [0.833; 0.845], model C1 C-index 0.838 [0.832; 0.844]). Similarly in the UK Biobank, the C-index did not the same and sensitivity increased after adding the interaction terms while specificity decreased. The sensitivity and specificity values between the two cohorts are different likely due to the well-known bias towards healthier individuals in the UK Biobank dataset (consistent with later analyses where the 7.5% risk threshold to classifies a smaller proportion of individuals into the high-risk group). However, the proportional gain in the sensitivity was consistent between the two cohorts (1.8% increase in HUNT2 and 1.2% in UK Biobank). **Figure 2** illustrates the effect of the Age*Sex*PGS_CAD_ term in the HUNT2 dataset. The hazard ratios (HRs) for the PGS_CAD_ on CAD 10-year risk with a model fit separately for males and females were not significantly different. However, we observed significant differences in model performance between the age groups when stratifying the dataset into three age-bins, demonstrating an age interaction. When further stratifying both males and females separately into age bins (approximating the Age*Sex*PGS_CAD_ interaction term in model C2), we observed small differences in the HRs between males and females in the same age bins. Simultaneously, we added an age*sex interaction term to ensure the model was valid by including all lower-level effects. The positive beta of this term indicates that irrespective of the PGS_CAD_, age increases the CAD risk more substantially for females compared to males, which could reflect the effect of menopause increasing the CAD risk in females(26).

**Table 1.** Diagnostic metrics for additive PGS_CAD_ model (model C1) and PGS_CAD_ model with all interactions (model C2) in HUNT2 and UK Biobank.

| Dataset | Risk model | Incident cases with risk $\geq 7.5\%$ | Incident cases with risk $< 7.5\%$ | Non-cases with risk $\geq 7.5\%$ | Non-cases with risk $< 7.5\%$ | Specificity/ Selectivity | Sensitivity/ Recall | C-index [95% confidence interval] |
| --- | --- | --- | --- | --- | --- | --- | --- | --- |
| HUNT2 | PGS<sub>CAD</sub> additive model (model C1) | 2290 | 684 | 11133 | 36929 | 76.8% | 77.0% | 0.838 [0.832; 0.844] |
| HUNT2 | PGS<sub>CAD</sub> with all interactions (model C2) | 2332 | 642 | 11539 | 36523 | 76.0% | 78.4% | 0.839 [0.833; 0.845] |
| UK Biobank | PGS<sub>CAD</sub> additive model (model C1) | 8688 | 8881 | 52201 | 233948 | 84.6% | 66.4% | 0.743 [0.739; 0.746] |
| UK Biobank | PGS<sub>CAD</sub> with all interactions (model C2) | 8996 | 8573 | 54913 | 231236 | 83.9% | 67.2% | 0.743 [0.740; 0.747] |

PGS<sub>CAD</sub>=Coronary artery disease polygenic score

**Figure 2.**
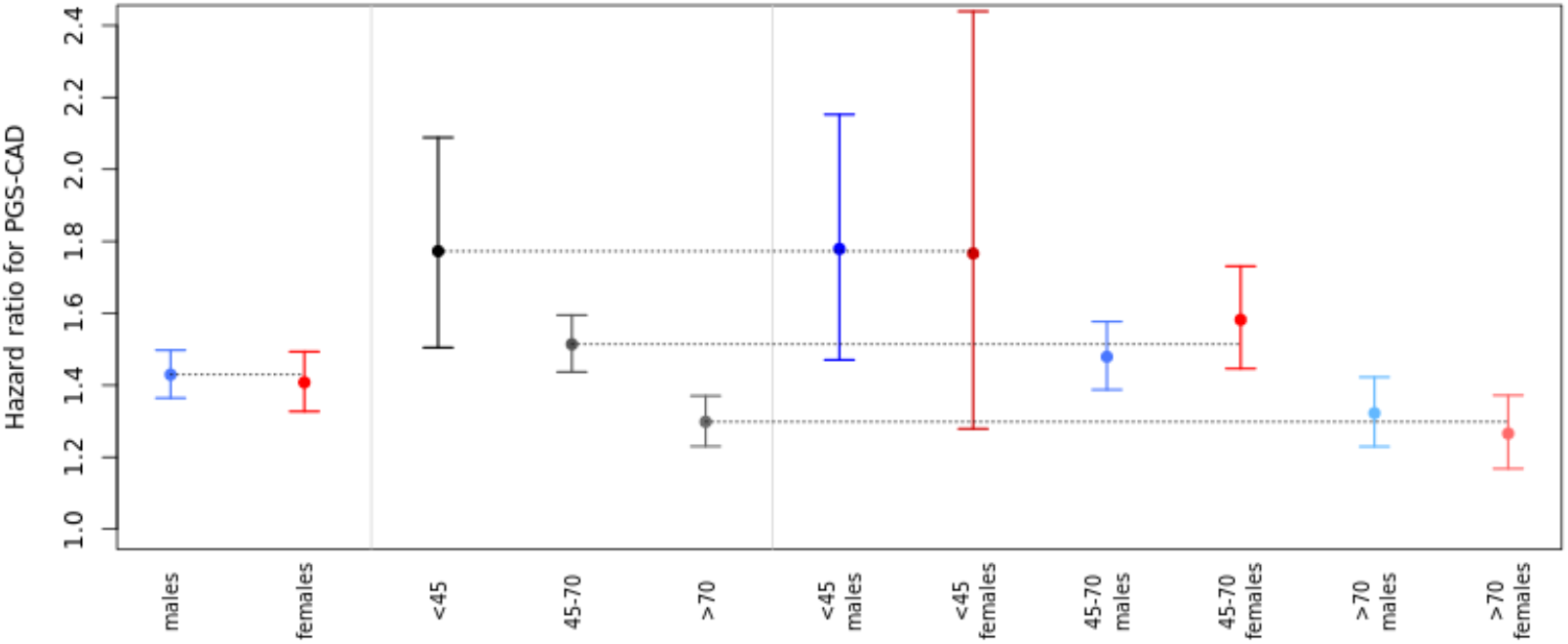
Age dependence of the sex-effect. This figure shows the hazard ratios for PGS_CAD_ in models fitted in 11 different subsets. Subsets were separated by sex (males, females), by age (<45-year-old, between 45 and 70, and more than 70-year-old) and finally stratified by both. All models have been adjusted for within-bin age and age^2^-effects and additionally the 3 age-bin models for sex.

### CAD Polygenic Score with ASCVD risk score

#### Joint Modeling

We expect that genetic risk will most likely be used in conjunction with or in addition to already existing risk estimates. With this in mind, we modeled the ASCVD risk score with the PGS_CAD_. Our model with additive effects only (model C3, **Supplemental Table 11**) had a higher C-index (0.842 [0.836; 0.848]) and slightly lower sensitivity than model C1 (76.8%), which suggests that including the ASCVD risk score (i.e., clinical score) on top of the PGS_CAD_ does not increase the number of identified cases, but rather affects the specificity, which increases from 76.8% (in model C1) to 77.4% (in model C3 which in includes ASCVD risk), and is observable by an increase in C-index. This finding could be caused by reduced transferability of PCE into Norwegian population. However, to evaluate the impact of the genetic interaction terms in a model with the clinical risk included, we tested the improvement in the model metrics by including the ASCVD risk score into the full PGS_CAD_ interaction model (model C4, **Table 2**). This model had the highest C-index (0.845 [0.839; 0.851]) and sensitivity (79.6%) of the combined prediction models. Moreover, when comparing the model with PGS_CAD_ and ASCVD predictors but without the interaction terms (model C3) to the same model but with full genetic interaction terms (model C4) the sensitivity increased from 76.8% to 79.7% while specificity decreased from 77.4% to 76.0%.

**Table 2.** Model statistics for PGS_CAD_ model with all interactions and ASCVD (model C4)

| **Variable/term**<br>**(units)** | **Effect** | **SE** | **HR** | **P-value** |
| --- | --- | --- | --- | --- |
| **Age<sup>2</sup> (year<sup>2</sup>)** | -2.8e-3 | 1.5e-4 | 0.997 | 9.4e-83 |
| **Sex (female=0, male=1)** | 3.24 | 0.255 | 25.47 | 7.1e-37 |
| **Age (year)** | 0.417 | 0.018 | 1.517 | 3.25e-119 |
| **PGSCAD (SD-unit)** | 1.21 | 0.182 | 3.369 | 2.3e-11 |
| **ASCVD (risk/100)** | 3.01 | 0.199 | 20.33 | 1.2e-51 |
| **Sex\*Age (year for males compared to females)** | -0.041 | 3.8e-3 | 0.960 | 7.3e-28 |
| **Sex\*PGSCAD (SD-units for males compared to females)** | -0.436 | 0.221 | 0.647 | 0.048 |
| **Age\*PGSCAD (SD-units per year)** | -0.013 | 2.6e-3 | 0.987 | 6.9e-7 |
| **Sex\*Age\*PGSCAD (SD-units per year for males compared to females)** | 6.2e-3 | 3.2e-3 | 1.006 | 0.052 |

SE=Standard error of the effect, HR=Hazard ratio, PGSCAD=Coronary artery disease polygenic score, SD=Standard deviation,
ASCVD=Atherosclerotic cardiovascular disease risk score

#### Two-step Approach

We also tested a scenario where the PGS_CAD_ could be added as an independent risk estimation tool to identify additional cases that were not already identified by their ASCVD risk score. This two-step case identification procedure is based on two sequential and independent risk estimates. After identifying high-risk individuals by the ASCVD risk score, we applied the PGS_CAD_ risk model to the remaining individuals, including interaction terms (effects coming from the model C2 for the full population with full population variable distributions). This staged approach, where ASCVD is first applied and PGS_CAD_ with genetic interaction terms is then applied, newly classified 3,235 individuals as high-risk (8.3% of the remaining dataset or 27.2% of the total dataset) totaling to 82.6% of the cases identified with the two-step approach. Among those newly classified, we observed 253 additional future cases during the 10-year follow-up (32.9% of the cases missed by the ASCVD score; **Figure 3**). If we used model C1 (the model without interaction terms) in the second step instead of the model C2 with the interactions, we would identify 81.5% of the total cases instead of 82.6%, highlighting the importance of interaction modeling also when using the sequential approach. The 253 additional incident cases identified using model C2 had a mean ASCVD risk of 4.73% ranging from 1.09% to 7.49%, suggesting the PGS_CAD_ provides information orthogonal to the ASCVD. We identified the same number of cases when applying the PGS_CAD_ model first (model C2) and then the ASCVD risk score (individuals that have either high PGS_CAD_ model C2 risk or high ASCVD risk). However, using the ASCVD first and then applying the genetic model may be more cost efficient as the number of samples needed to be genotyped is lower (only those with low ASCVD risk), and follows the current standard clinical practice for the first stage.

**Figure 3.**
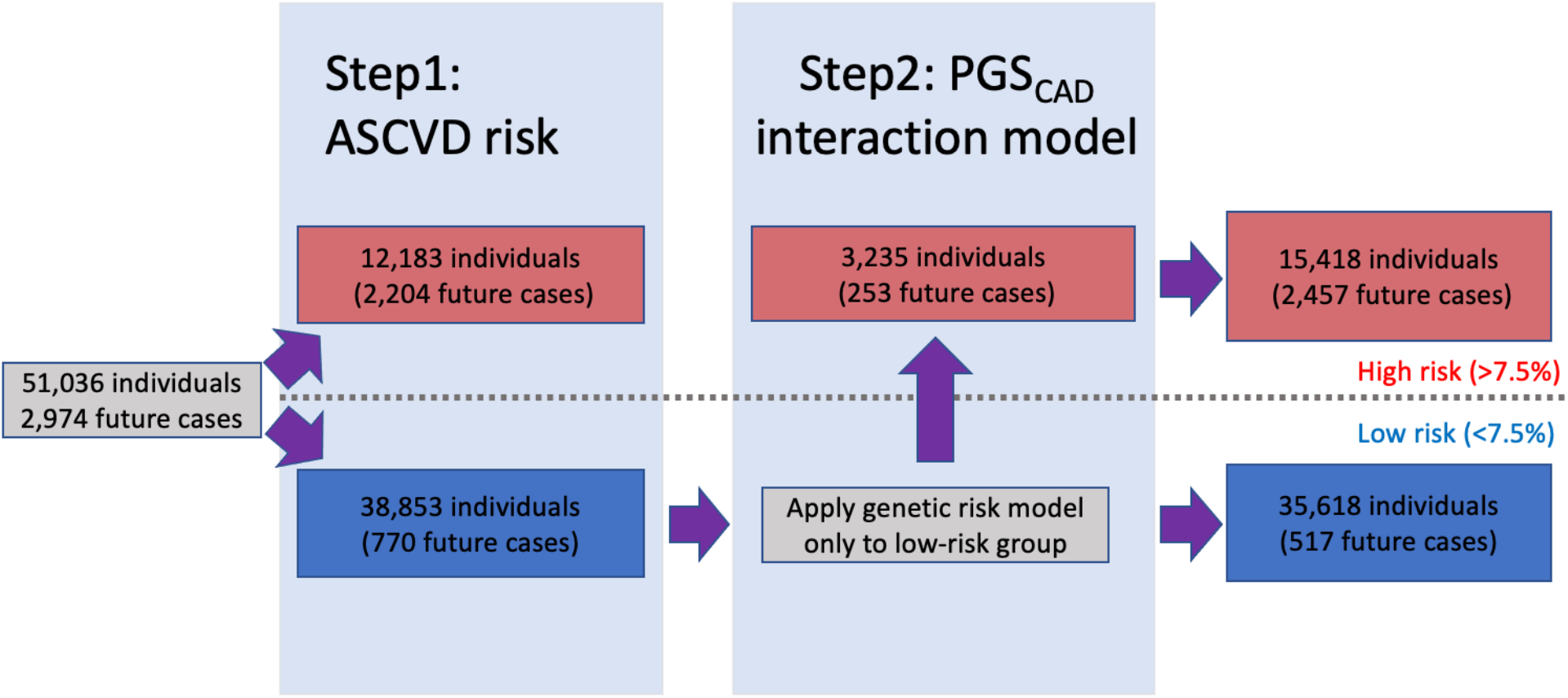
Illustration of the two-step approach combining ASCVD risk and the genetic risk model. This figure shows how combining the ASCVD and the genetic risk model with interactions in two consecutive steps allows for identification of additional cases.

### Sex-Specific Models and Sex-Specificity of Model Metrics

The currently applied clinical risk scores are typically applied to males and females separately instead of using sex-interaction models. To test the applicability of our PGS_CAD_ interaction models in the similar manner, we tested the performance of models allowing for age-dependence of the PGS_CAD_ separately in males and females. First, we evaluated the PGS_CAD_ model without interactions (model S1, **Supplemental Tables 12A-B**). The C-indexes observed were 0.850 [0.840; 0.860] for females and 0.816 [0.808; 0.824] for males, and the magnitude of the HR for the PGS_CAD_ was similar for both sexes (HR females = 1.41 [1.33; 1.49], HR males = 1.43 [1.37; 1.50]).

The inclusion of the PGS_CAD_*Age interaction into the models (**Supplemental Tables 13A-B**) did not notably change the C-indexes, even though the interaction term was significant for both sexes (P-value in females = 1.85e-6, in males = 8.91e-4). However, the sensitivity increased for both sexes in HUNT2 (**Table 3A-B**). In UK Biobank the sensitivity only increased for males (**Supplementary Tables 14A-B**). Finally, both C-index and sensitivity increased for both males and females when adding the ASCVD risk score to the model (model S3, **Supplemental Tables 15A-B**). Lastly, we performed the two-step process described earlier for males and females separately by including i). the conventional ASCVD risk and ii) PGS_CAD_ with age-interaction term. Using the two-step approach, we correctly re-classified an additional 194 and 59 future cases for males and females, respectively (38.3% and 22.4% of the cases missed by the ASCVD risk assessment). We observed increased sensitivity by the two-step approach also in the UK Biobank. The corresponding numbers without the interaction terms in the second step were 183 future cases for males and 51 for females (36.1% and 19.4% of the cases missed by the ASCVD risk score).

**Table 3A-B.**
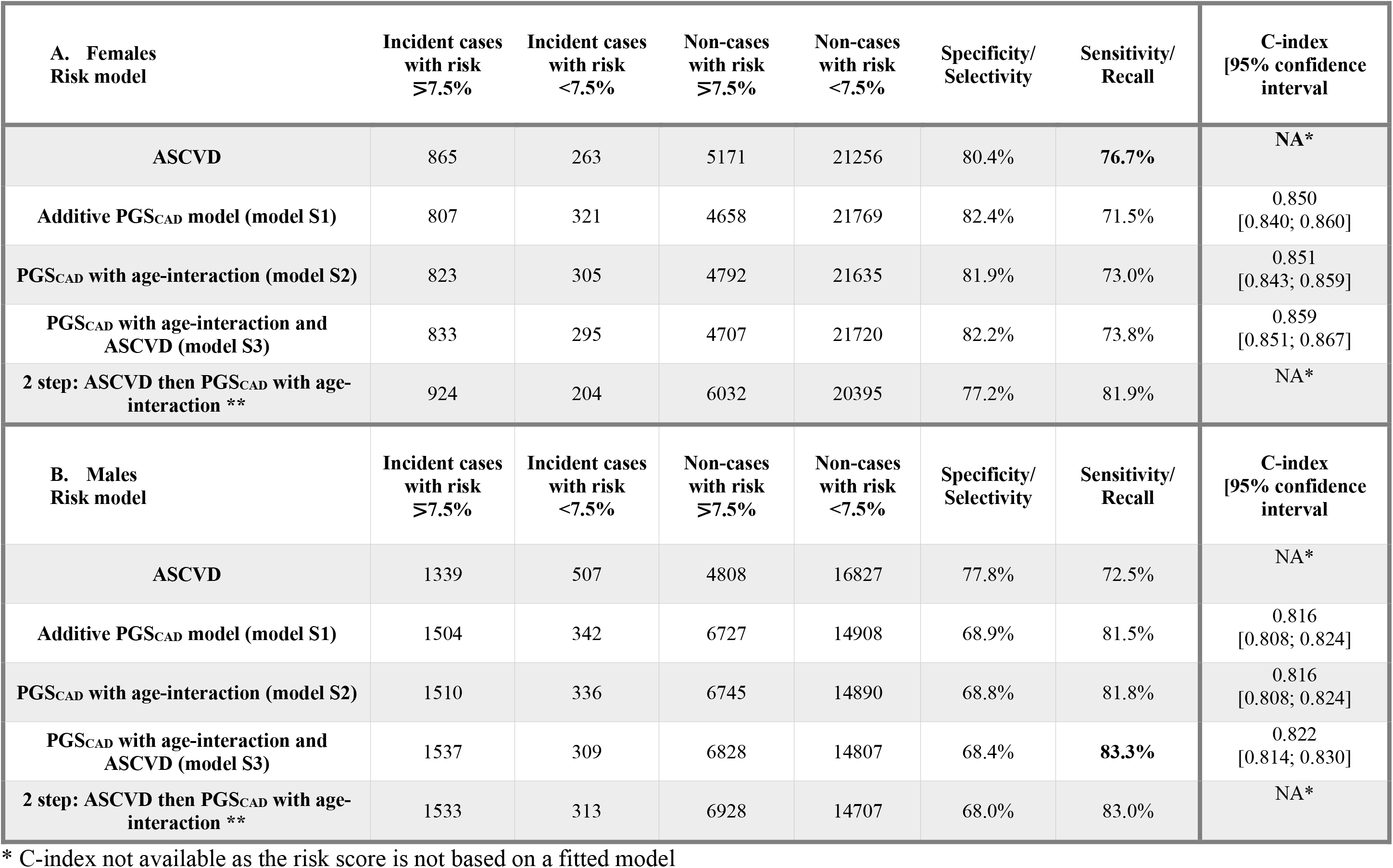
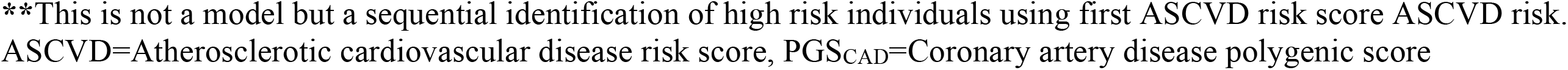
Diagnostic metrics for sex-stratified models and ASCVD risk score.

| **A. Females**<br>**Risk model** | **Incident cases**<br>**with risk**<br>**≥7.5%** | **Incident cases**<br>**with risk**<br>**<7.5%** | **Non-cases**<br>**with risk**<br>**≥7.5%** | **Non-cases**<br>**with risk**<br>**<7.5%** | **Specificity/**<br>**Selectivity** | **Sensitivity/**<br>**Recall** | **C-index**<br>**[95% confidence**<br>**interval]** |
| --- | --- | --- | --- | --- | --- | --- | --- |
| **ASCVD** | 865 | 263 | 5171 | 21256 | 80.4% | **76.7%** | NA* |
| **Additive PGSCAD model (model S1)** | 807 | 321 | 4658 | 21769 | 82.4% | 71.5% | 0.850<br>[0.840; 0.860] |
| **PGSCAD with age-interaction (model S2)** | 823 | 305 | 4792 | 21635 | 81.9% | 73.0% | 0.851<br>[0.843; 0.859] |
| **PGSCAD with age-interaction and**<br>**ASCVD (model S3)** | 833 | 295 | 4707 | 21720 | 82.2% | 73.8% | 0.859<br>[0.851; 0.867] |
| **2 step: ASCVD then PGSCAD with age-**<br>**interaction \*\*** | 924 | 204 | 6032 | 20395 | 77.2% | 81.9% | NA* |
| **B. Males**<br>**Risk model** | **Incident cases**<br>**with risk**<br>**≥7.5%** | **Incident cases**<br>**with risk**<br>**<7.5%** | **Non-cases**<br>**with risk**<br>**≥7.5%** | **Non-cases**<br>**with risk**<br>**<7.5%** | **Specificity/**<br>**Selectivity** | **Sensitivity/**<br>**Recall** | **C-index**<br>**[95% confidence**<br>**interval]** |
| **ASCVD** | 1339 | 507 | 4808 | 16827 | 77.8% | 72.5% | NA* |
| **Additive PGSCAD model (model S1)** | 1504 | 342 | 6727 | 14908 | 68.9% | 81.5% | 0.816<br>[0.808; 0.824] |
| **PGSCAD with age-interaction (model S2)** | 1510 | 336 | 6745 | 14890 | 68.8% | 81.8% | 0.816<br>[0.808; 0.824] |
| **PGSCAD with age-interaction and**<br>**ASCVD (model S3)** | 1537 | 309 | 6828 | 14807 | 68.4% | **83.3%** | 0.822<br>[0.814; 0.830] |
| **2 step: ASCVD then PGSCAD with age-**<br>**interaction \*\*** | 1533 | 313 | 6928 | 14707 | 68.0% | 83.0% | NA* |

\* C-index not available as the risk score is not based on a fitted model
**\*\*This is not a model but a sequential identification of high risk individuals using first ASCVD risk score ASCVD risk.** ASCVD=Atherosclerotic cardiovascular disease risk score, PGS<sub>CAD</sub>=Coronary artery disease polygenic score

### Sex-Specific Model Metrics and the Effect of the Risk Threshold

We saw lower sensitivity and higher specificity for females compared to males in all sex stratified models that included genetic information. These two metrics are dependent on the risk-threshold. Therefore, we tested how changing the threshold would affect the risk classification. The sensitivity and specificity for males and females for varied risk-thresholds are presented in **Supplemental Figures 4-7**. For all of the stratified models, the percentages of individuals in the high-risk group were higher for males than for females at any given risk threshold. This finding was expected given that females have a lower overall prevalence of CAD. The proportion of individuals in the high-risk group based on ASCVD risk was close between the two sexes (**Supplemental Figure 7**). This is most likely due to the underestimation of the ASCVD risk seen for males when applying the ASCVD risk calculation to our test dataset (**Supplemental Figure 8**). **Supplemental Figure 9** shows the risk calibration for the model S3 as comparison.

However, lower sensitivity was observed for females for models that include the PGS_CAD_. To achieve the same sensitivity observed for males at the 7.5% risk threshold (81.4%), we would need to lower the risk threshold in females to 5.0% (**Supplemental Figures 4,5 and 6**). In all three models (S1-3), the specificity in females with the 5.0% risk threshold was better than the specificity in males with the 7.5% risk threshold.

## DISCUSSION

This study evaluated several statistical approaches in two population-based datasets to fine-tune the prediction of individuals at risk for 10-year CAD events by accounting for different rates and age distributions of cardiovascular disease in males and females. We found that the C-index and sensitivity of the 10-year prediction of CAD improved by including sex and age interactions when modeling PGS_CAD_ compared to a model without interaction effects in both datasets. Inclusion of the interaction terms most likely corrects for the survival bias observed and the implications of these results highlight the importance of modeling age and sex interactions in predicting CAD events with genetic information.

In our baseline correlation checks, we observed significant associations between the genetic score and both age and sex, and replicated these findings in the UK Biobank. The observed associations suggest non-random selection related to genetics in the study cohorts, and we contend that the age association is derived from the survival bias of individuals with lower genetic risk of CAD. The sex association is most likely derived from the earlier onset of CAD in males, which enhances the survival bias of those with lower genetic risk in males. We expect these biases to be present in all cross-sectional studies where the age ranges over the expected age-of-onset of the studied disease. Moreover, the same biases are most likely also present in populations where risk estimates are applied to identify high-risk individuals.

We evaluated the potential incorporation of genetic information into identifying at-risk individuals by applying joint modeling or by applying two risk estimates (clinical and genetic) in a sequential manner. Both of these approaches showed increased number of cases identified when the age and sex dependent behavior of the genetic risk was taken into account. Additionally, with genome-wide genotyping being translated into clinical settings, CAD risk prediction may be enhanced by the sequential two-step approach we evaluate here: i.) first apply the existing clinical score (i.e., PCE/ASCVD risk score) and ii.) from those identified with a low ASCVD risk, apply a second model incorporating age, sex, and genetic information with age and sex interactions to identify additional high-risk individuals (**Figure 4**). Using our two-step approach with a set risk threshold of 7.5%, we identified a total of 82.6% of incident CAD diagnoses (74.1% by ASCVD risk estimation and an additional 8.5% by the PGS_CAD_ interaction model). The newly identified future cases in the second step suggests that incorporating genetic information including age and sex interaction modeling captures cases that do not yet show clinical signs of atherosclerosis or hypertension (which are the biggest clinical contributors to the ASCVD risk after age and sex). The implications of these results could be two-fold i.) clinicians maintain the ability to identify high-risk individuals using the ASCVD risk tool, and ii.) clinicians with access to genetic information on patients are then able to more accurately discern which additional individuals may benefit from timely prevention strategies (**Central Illustration**). Implementation of this approach will require a large study with diverse populations to tests risk factors including genetic information to ascertain population level effects that can be applied to a single patient in clinical practice.

**Figure 4.**
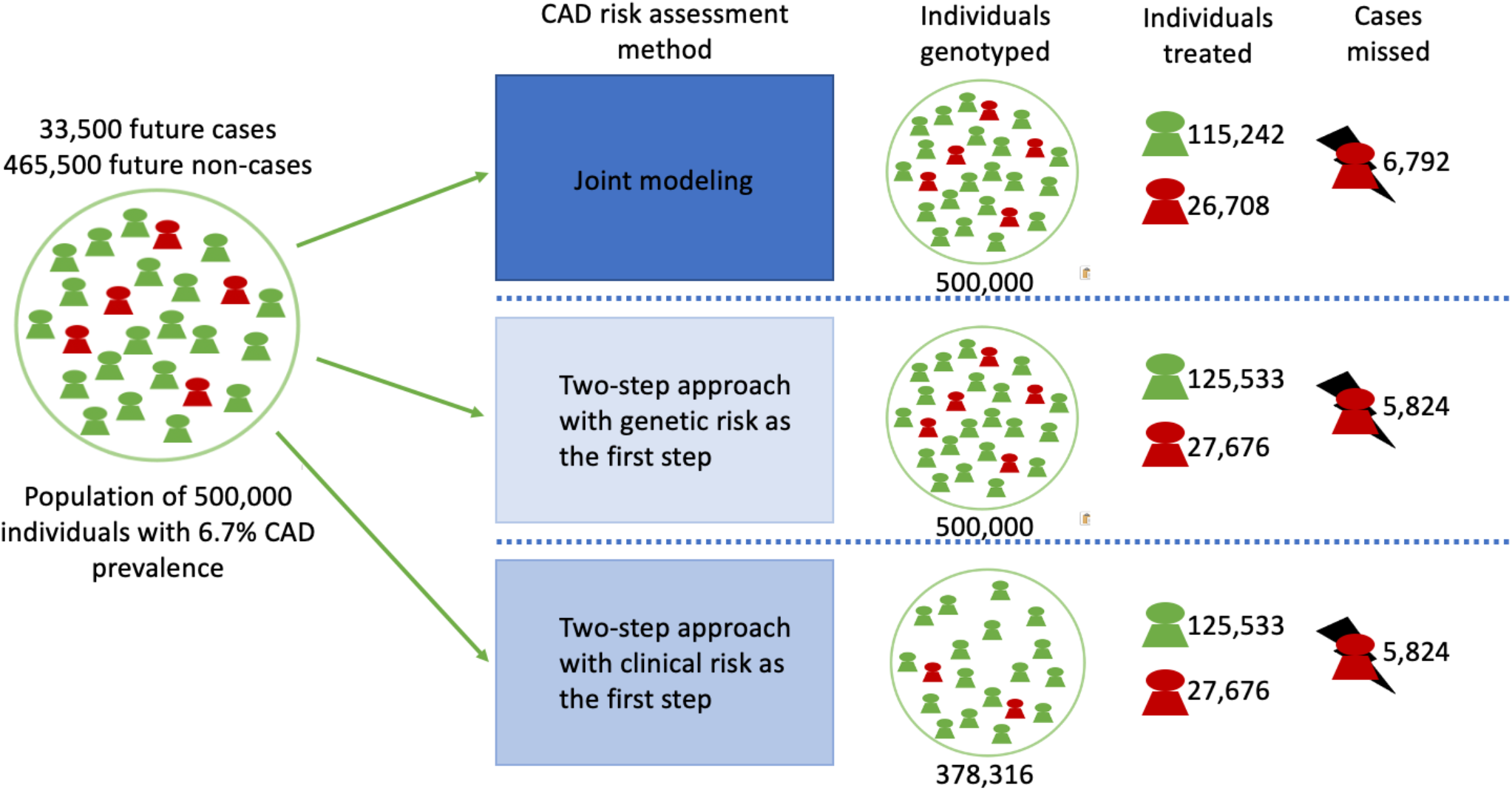
Demonstration of the different risk models implementing clinical and genetic risk with interactions in a hypothetical population of 500,000 people. The CAD prevalence used in the demonstration is based on the current CAD prevalence in the US.

For both cohorts, the sensitivity for females was consistently lower than for males. In the HUNT2 dataset, we found that similar sensitivity to predict female cases could be achieved by lowering the risk threshold for preventive therapies from 7.5% to 5.0%. Additionally, this would not result in a higher proportion of females recommended for treatment relative to males. We suggest that the risk threshold used in the genetic screening should be independently evaluated in males and females before applying genetic information in an equal manner in the clinical setting. For example, in our dataset, if we changed the risk threshold from 7.5% to 5.0% in females when applying the two-step sequential approach, we would increase the identification of cases from 81.9% to 86.2%% without increasing the proportional amount of females suggested for treatment (25.3%) relative to males recommended for treatment (36.0%).

Our study has important limitations. First, the datasets used in this study, HUNT2 and UK Biobank, are sampled from different populations than the datasets in which the ASCVD score was originally created (different ancestry, country of residence, younger, and healthier). Moreover, the ASCVD score was developed to evaluate the risk of developing CAD or stroke. In our study, we used the ASCVD score to predict CAD event or death during the 10-year follow-up time. This approach may have caused the miscalibration observed in the HUNT2 study, which limits our ability to perform unbiased one-to-one comparisons between the performance of these scoring methods. However, the trends and conclusions reported herein do not rely on the exact ASCVD risk, but rather, compare the change in the metrics when modeling genetics with and without age and sex interaction terms. Second, the participants in this study are of European ancestry, and therefore, the results may not be generalizable to populations with other ancestries (27). Additional studies are needed to determine the importance of interaction effects in the genetic prediction of other traits and in diverse populations with different rates of clinical risk factors such as hypertension and high LDL cholesterol. Third, we tested the performance of the interaction models against only one clinical score, albeit the one recommended by the American Heart Association(2). Lastly, our models were based on only a single PGS, although the performance of several different genome-wide PGSs (i.e. those derived from statistical methods such as metaGRS, LDpred or PRS-CS) have shown to be nearly equivalent in CAD prediction(28).

### Conclusion

All populations screened for CAD risk are subject to survival bias that shows as a depletion of high PGS individuals. Therefore, we suggest using age and sex interactions with the PGS in disease prediction. To predict future CAD events, the best performing models we identified utilize both clinical and genetic information including interactions -- whether applied as a single model or in a sequential two-step process. Moreover, CAD prediction studies with genetic information should focus on the sex-specific behavior of the predictors and prediction models to account for sex-specific genetic effects and differences in the incidence of CAD events between males and females.

### Perspective

#### Competency in Patient Care

Our results highlight the importance and complexity of the genetic risk in the predicting CAD events and suggest including age and sex interactions in prediction models to identify more high-risk individuals for early prevention, in addition to existing clinical tools. Application of polygenic risk scores to guide early preventive therapy needs to be considered in the context that risk estimation differs based on the age and sex of the patient.

#### Translational Outlook

Our findings present a path forward for future studies to comprehensively evaluate the age and sex-specific impact of risk-predicting genetic information. Fine-tuning the risk threshold for males and females separately is required to provide optimized risk information to patients and guide clinical decision-making focused on prevention.

## Data Availability

UK Biobank is freely available for research purposes (https://www.ukbiobank.ac.uk). HUNT2 summary level data is available upon reasonable request.

## Acknowledgements

The authors thank the HUNT and UK Biobank participants for their contributions to research. HUNT-MI study, which comprises the genetic investigations of the HUNT Study, is a collaboration between investigators from the HUNT study and University of Michigan Medical School and the University of Michigan School of Public Health. The K.G. Jebsen Center for Genetic Epidemiology is financed by Stiftelsen Kristian Gerhard Jebsen; Faculty of Medicine and Health Sciences, NTNU, Norwegian University of Science and Technology (NTNU) and Central Norway Regional Health Authority. This research has been conducted using the UK Biobank Resource under Application Number 7439. This work was supported by core funding from the: British Heart Foundation (RG/13/13/30194; RG/18/13/33946), Cambridge BHF Centre of Research Excellence (RE/13/6/30180) and NIHR Cambridge Biomedical Research Centre (BRC-1215-20014) [The views expressed are those of the author(s) and not necessarily those of the NIHR or the Department of Health and Social Care]. This work was also supported by Health Data Research UK, which is funded by the UK Medical Research Council, Engineering and Physical Sciences Research Council, Economic and Social Research Council, Department of Health and Social Care (England), Chief Scientist Office of the Scottish Government Health and Social Care Directorates, Health and Social Care Research and Development Division (Welsh Government), Public Health Agency (Northern Ireland), British Heart Foundation and Wellcome. The authors thank Kuan-Han Wu for his important graphical contributions.

## Abbreviations

(ASCVD): Atherosclerotic cardiovascular disease
(Harrell’s C-index, referred to as C-index throughout the study): Concordance index
(CAD): Coronary artery disease
(HRs): Hazard ratios
(PCE): Pooled Cohort Equation
(PGS): polygenic score
(HUNT): Trøndelag Health Study
(UK Biobank): United Kingdom Biobank

## Author Information

I.S., K.H and C.J.W. designed the study. I.S., S.C.R and B.N.W. analyzed the data. A.H.S., M.E.G. and L.T. contributed to the phenotype harmonization. N.R.S. and W.E.H. provided clinical expertise. I.S., W.E.H., S.C.R., M.I., K.H. and C.J.W. wrote the paper. All the authors read and revised the manuscript.

## Ethics Declaration

Participation in the HUNT Study is based on informed consent and the study has been approved by the Data Inspectorate and the Regional Ethics Committee for Medical Research in Norway (REK: 2014/144)

## Central Illustration

**Coronary Artery Disease Polygenic Scores Used for Risk Prediction Exhibit Age and Sex-Interactions**

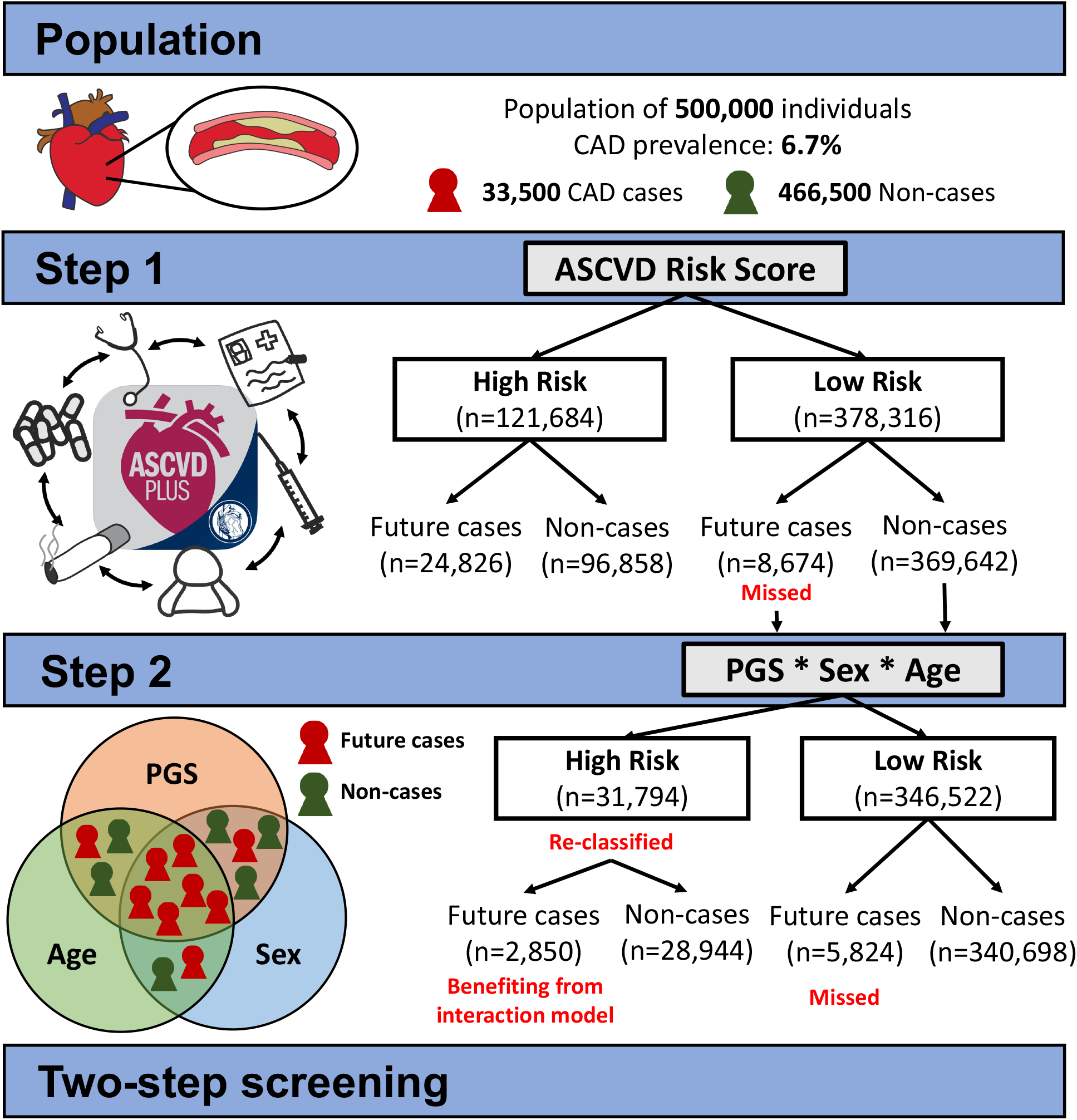

Using the two step approach we were able to identify a total of 82.6% of incident CAD cases; 74.1% by ASCVD risk score and an additional 8.5% by the PGSCAD interaction model. These results highlight the importance of combined utilization of both traditional and genetic risk estimation as well as the implementation of the age and sex dependent behavior of the genetic risk.

