## Supplementary Material for "Sex-specific survival bias and interaction modeling in coronary artery disease risk prediction"

### Supplemental Information

#### Supplemental Methods

##### Phenotype definitions in HUNT2 dataset

HUNT2 hospital registry and cause of death data was linked to the genotype data. The cause of death registry data was collected at the end of November 2017 and the hospital registry data at the end of December 2017. The end of the follow-up was therefore set at the end of November 2017. Using these two registries, the CAD endpoint was defined as the first occurrence of any of the following ICD-codes in either of hospital or death registry: myocardial infarction or acute ischemic heart disease (ICD10: I21-I24; ICD9: 410), or chronic ischemic heart disease (ICD-10: I25 excluding I25.3, I.25; ICD9: 411, 412, 414.0, 414.8, 414.9). This definition of CAD follows the definition of intermediate CAD by Nikpay et al.(25) with the exception of not having the procedure codes available. At baseline, blood pressure medication was collected by self-report, and prevalent diabetes was defined by a combination of self-report and hospital registry data (ICD10: E10-E11 excluding E11.5, E13-E13.9 excluding E13.3, E14.9; ICD9: 250-250.99).

##### Phenotype definitions in UK Biobank dataset

Incident CAD defined as for intermediate CAD in CARDIoGRAMplusC4D (25). Follow-up is determined as time to first event from baseline, truncating follow-up at 1st February 2020 to avoid confounding from COVID-19 pandemic (either due to SARS-CoV2 exposure or behavioural changes/changes to environment due to lockdown).

- ICD-10: I21-I24, I25.1, I25.2, I25.5-I25.9
- OPSC-4: K40-K46, K49, K50.1, K50.2, K50.4, K75

Prevalent CAD definitions include the above, and additionally:

- ICD-9: 410-412, 414.0, 414.8, 414.9

- (All events with ICD-9 coding are retrospective in UKBB)

- Self-reported heart attack/myocardial infarction at verbal interview (UKBB field #20002, code 1075)
- Self-reported coronary angioplasty (ptca) +/- stent, coronary artery bypass grafts (cabg), or triple heart bypass at verbal interview (UKBB field #20004, codes 1070, 1095, and 1523)
- Self-reported heart attack at touchscreen interview (UKBB field #6150)

Note different nations (England/Wales/Scotland) have different maximum follow-up dates. While hospital records in England and Scotland go beyond 1$^{st}$ Feb 2020, hospital records in Wales are currently only available up until 6$^{th}$ March 2018. In the whole of UKBB (no sample criteria applied), 4.1% of participants have maximum follow-up in the Welsh hospital system. Scotland also has a higher median follow-up time, as on average baseline assessment happened earlier in Scotland than in England or Wales.

### Supplemental Tables

**Supplemental Table 1. Cohort descriptives of HUNT2 dataset.** Baseline characteristics of HUNT2

| Variable (units) | Males (N=23,481) Mean (SD) / N (Proportion) | Females (N=27,555) Mean (SD) / N (Proportion) | Incident case males (N=1,846) Mean (SD) / N (Proportion) | Incident case females (N=1,128) Mean (SD) / N (Proportion) | Non-case males (N=21,635) Mean (SD) / N (Proportion) | Non-case females (N=26,427) Mean (SD) / N (Proportion) |
|---|---|---|---|---|---|---|
| **Age (years)** | 48.7 (16.1) | 49.0 (16.5) | 65.3 (12.0) | 69.2 (11.5) | 47.3 (15.6) | 48.1 (16.1) |
| **Total cholesterol (mmol/l)** | 5.77 (1.13) | 5.89 (1.31) | 6.37 (1.11) | 6.89 (1.26) | 5.71 (1.12) | 5.85 (1.29) |
| **HDL cholesterol (mmol/l)** | 1.26 (0.33) | 1.51 (0.38) | 1.20 (0.33) | 1.42 (0.39) | 1.26 (0.33) | 1.51 (0.38) |
| **Systolic Blood pressure (mm Hg)** | 139.34 (18.55) | 134.56 (22.84) | 150.89 (22.61) | 157.62 (25.17) | 138.36 (17.82) | 133.58 (22.20) |
| **Currently Smoking (yes/no)** | 6754 (28.8%) | 8278 (30.0%) | 653 (35.4%) | 327 (29.0%) | 6101 (28.2%) | 7951 (30.1%) |
| **Blood Pressure medication (yes/no)** | 2459 (10.5%) | 3622 (13.1%) | 513 (27.8%) | 472 (41.8%) | 1946 (9.0%) | 3150 (11.9%) |
| **Prevalent Diabetes (yes/no)** | 659 (2.8%) | 719 (2.6%) | 171 (9.3%) | 133 (11.8%) | 488 (2.3%) | 586 (2.2%) |

N=Number of samples, SD=standard deviation

**Supplemental Table 2. Cohort descriptives of UK Biobank dataset.** Baseline characteristics of the UK Biobank

| Variable (units) | Males (N= 157,570) Mean (SD) / N (Proportion) | Females (N= 214,840) Mean (SD) / N (Proportion) | Incident case males (N= 11,093) Mean (SD) / N (Proportion) | Incident case females (N= 6,476) Mean (SD) / N (Proportion) | Non-case males (N= 146,477) Mean (SD) / N (Proportion) | Non-case females (N= 208,364) Mean (SD) / N (Proportion) |
|---|---|---|---|---|---|---|
| **Age (years)** | 56.1 (8.15) | 56.3 (7.93) | 60.0 (6.98) | 61.0 (6.59) | 55.8 (8.16) | 56.2 (7.93) |
| **Total cholesterol (mmol/l)** | 5.77 (1.02) | 6.02 (1.08) | 5.93 (1.08) | 6.33 (1.14) | 5.76 (1.01) | 6.01 (1.08) |
| **HDL cholesterol (mmol/l)** | 1.31 (0.31) | 1.62 (0.38) | 1.26 (0.31) | 1.53 (0.37) | 1.31 (0.31) | 1.62 (0.37) |
| **Systolic Blood pressure (mm Hg)** | 140.42 (17.34) | 134.33 (19.13) | 145.79 (18.28) | 142.45 (19.87) | 140.01 (17.20) | 134.07 (19.05) |
| **Currently Smoking (yes/no)** | 19,301 (12.3%) | 18,712 (8.71%) | 1,810 (16.3%) | 949 (14.7%) | 17,491 (11.9%) | 17,763 (8.53%) |
| **Blood Pressure medication (yes/no)** | 20,013 (12.7%) | 24,593 (11.4%) | 2,471 (22.3%) | 1,627 (25.1%) | 17,542 (12.0%) | 22,966 (11.0%) |
| **Prevalent Diabetes (yes/no)** | 2,769 (1.76%) | 1,760 (0.82%) | 430 (3.89%) | 179 (2.77%) | 2,339 (1.60%) | 1,581 (0.76%) |

N=Number of samples, SD=standard deviation

**Supplemental Table 3. Summary of the used Cox Proportional Hazards models**

| Model name | Variables in the linear predictor | Applied dataset |
|---|---|---|
| **$PGS_{CAD}$ additive model (model C1)** | Age+Age$^2$+Sex+$PGS_{CAD}$ | Combined |
| **$PGS_{CAD}$ with all interactions (model C2)** | Age+Age$^2$+Sex+$PGS_{CAD}$+Sex*$PGS_{CAD}$+Age*$PGS_{CAD}$+ Sex*Age +Sex*Age*PGSCAD | Combined |
| **Additive $PGS_{CAD}$ with ASCVD (model C3)** | Age+Age$^2$+Sex+$PGS_{CAD}$+ASCVD | Combined |
| **$PGS_{CAD}$ with all interactions and ASCVD (model C4)** | Age+Age$^2$+Sex+$PGS_{CAD}$+Sex*$PGS_{CAD}$+Age*$PGS_{CAD}$+ Sex*Age +Sex*Age*PGSCAD +ASCVD | Combined |
| **Additive $PGS_{CAD}$ model (model S1)** | Age+Age$^2$+$PGS_{CAD}$ | Sex stratified |
| **$PGS_{CAD}$ with age-interaction (model S2)** | Age+Age$^2$+$PGS_{CAD}$+Age*$PGS_{CAD}$ | Sex stratified |
| **$PGS_{CAD}$ with age-interaction and ASCVD (model S3)** | Age+Age$^2$+$PGS_{CAD}$+Age*$PGS_{CAD}$+ASCVD | Sex stratified |

ASCVD=Atherosclerotic cardiovascular disease risk score,
$PGS_{CAD}$=Coronary artery disease polygenic score

**Supplemental Table 4A-E. Baseline linear regression models in HUNT2 dataset**

| A. **$PGS_{CAD}$~Sex** | | | |
|---|---|---|---|
| **Variable** | **Beta** | **SE** | **P-value** |
| **Intercept** | 0.015 | 0.006 | 0.013 |
| **Sex (0=female, 1=male)** | -0.056 | 0.009 | 3.0e-10 |
| B. **$PGS_{CAD}$~Age** | | | |
| **Intercept** | 0.136 | 0.014 | <2.22e-16 |
| **Age (year)** | -0.003 | 2.7e-4 | <2.22e-16 |
| C. **Males only $PGS_{CAD}$~Age** | | | |
| **Intercept** | 0.114 | 0.021 | 4.2e-8e-9 |
| **Age (year)** | -3.2e-3 | 4.0e-4 | 4.4e-15 |
| D. **Females only $PGS_{CAD}$~Age** | | | |
| **Intercept** | 0.137 | 0.019 | 3.4e-13 |
| **Age (year)** | -2.5e-3 | 3.6e-4 | 7.8e-12 |
| E. **$PGS_{CAD}$~Sex+Age+Sex*Age** | | | |
| **Intercept** | 0.137 | 0.019 | 3.4e-13 |
| **Age (year)** | -2.5e-3 | 3.6e-4 | 7.9e-12 |
| **Sex (0=female, 1=male)** | -0.023 | 0.028 | 0.408 |
| **Sex*Age (year for males compared to females)** | -6.8e-4 | 5.4e-4 | 0.210 |

$PGS_{CAD}$=Coronary artery disease polygenic score

**Supplemental Table 5. Replication of the baseline $PGS_{CAD}$ trends in UKBB.**

| | | | |
|---|---|---|---|
| A. **$PGS_{CAD}$~Sex** | | | |
| **Variable** | **Beta** | **SE** | **P-value** |
| **Intercept** | 0.052 | 0.006 | 2e-20 |
| **Sex (0=female,1=male)** | -0.034 | 0.003 | 3e-24 |
| B. **$PGS_{CAD}$~Age** | | | |
| **Intercept** | 0.27 | 0.013 | 5e-103 |
| **Age (year)** | -0.004 | 0.000 | 8e-94 |
| C. **Males only $PGS_{CAD}$~Age** | | | |
| **Intercept** | 0.35 | 0.019 | 8e-76 |
| **Age (year)** | -0.006 | 0.000 | 2e-72 |
| D. **Females only $PGS_{CAD}$~Age** | | | |
| **Intercept** | 0.21 | 0.017 | 1e-36 |
| **Age (year)** | -0.003 | 0.000 | 1e-31 |
| E. **$PGS_{CAD}$~Sex+Age+Sex*Age** | | | |
| **Intercept** | 0.23 | 0.016 | 6e-46 |
| **Age (year)** | -0.003 | 0.000 | 6e-32 |
| **Sex (0=female, 1=male)** | 0.097 | 0.023 | 3e-5 |
| **Sex*Age (year for males compared to females)** | -0.002 | 0.000 | 1e-8 |

$PGS_{CAD}$=Coronary artery disease polygenic score

**Supplemental Table 6A-B. Baseline linear regression models with all variables in HUNT2 (A) and UKBB (B) datasets with prevalent cases in HUNT2 (N=52,491) and prevalent cases and baseline statin users in UKBB (N=456,702)**

| A. **$PGS_{CAD}$~Sex+Age+Sex*Age in HUNT2 including prevalent cases** | | | |
|---|---|---|---|
| **Variable** | **Beta** | **SE** | **P-value** |
| **Intercept** | 0.118 | 0.019 | 3.6e-10 |
| **Age (year)** | -2.0e-3 | 3.6e-4 | 5.7e-8 |
| **Sex (0=female, 1=male)** | -0.035 | 0.028 | 0.210 |
| **Sex*Age (year for males compared to females)** | -2.0e-4 | 5.3e-4 | 0.705 |
| B. **$PGS_{CAD}$~Sex+Age+Sex*Age in UKBB including prevalent cases** | | | |
| **Intercept** | 0.084 | 0.015 | 4e-8 |
| **Age (year)** | -0.001 | 0.000 | 0.004 |
| **Sex (0=female, 1=male)** | 0.030 | 0.021 | 0.16 |
| **Sex*Age (year for males compared to females)** | -0.001 | 0.000 | 0.12 |

$PGS_{CAD}$=Coronary artery disease polygenic score

**Supplemental Table 8. Cox proportional hazards model statistics for the additive $PGS_{CAD}$ model (model C1)**

| Variable/term (units) | Effect | SE | HR | P-value |
|---|---|---|---|---|
| $Age^2$ ($year^2$) | -1.27e-3 | 9.9e-5 | 0.999 | 1.02e-37 |
| Sex (female=0, male=1) | 0.774 | 0.038 | 2.17 | 8.52e-93 |
| Age (year) | 0.237 | 0.013 | 1.27 | 2.63e-77 |
| $PGS_{CAD}$ (SD-units) | 0.354 | 0.019 | 1.42 | 3.08e-80 |

SE=Standard error of the effect, HR=Hazard ratio, $PGS_{CAD}$=Coronary artery disease polygenic score, SD=Standard deviation

**Supplemental Table 9. Diagnostic metrics for Pooled cohort equation and all models with the CAD polygenic score in HUNT2 dataset.**

| Risk model | Incident cases with risk ⩾7.5% | Incident cases with risk <7.5% | Non-cases with risk ⩾7.5% | Non-cases with risk <7.5% | Specificity/ Selectivity | Sensitivity/ Recall | C-index [95% confidence interval] |
|---|---|---|---|---|---|---|---|
| ASCVD risk score | 2204 | 770 | 9979 | 38083 | 79.2% | 74.1% | NA* |
| $PGS_{CAD}$ additive model (model C1) | 2290 | 684 | 11127 | 36935 | 76.8% | 77.0% | 0.838 [0.832; 0.844] |
| $PGS_{CAD}$ with all interactions (model C2) | 2333 | 641 | 11536 | 36526 | 76.0% | 78.4% | 0.839 [0.833; 0.845] |
| $PGS_{CAD}$ additive with ASCVD (model C3) | 2284 | 690 | 10866 | 37196 | 77.4% | 76.8% | 0.842 [0.836; 0.848] |
| $PGS_{CAD}$ with interactions and ASCVD (model C4) | 2371 | 603 | 11553 | 36509 | 76.0% | **79.7%** | 0.845 [0.839; 0.851] |
| 2. step: ASCVD then $PGS_{CAD}$ with interactions* | 2457 | 517 | 12960 | 35102 | 73.0% | 82.6% | NA* |

* C-index not available as the risk score is not based on a fitted model
**This is not a model but a sequential identification of high risk individuals using first ASCVD risk score and then the model with $PGS_{CAD}$ with both interactions within individuals with low ASCVD risk.
ASCVD=Atherosclerotic cardiovascular disease risk score,
$PGS_{CAD}$=Coronary artery disease polygenic score

**Supplemental Table 10. Model statistics for $PGS_{CAD}$ model with all interactions (model C2)**

| Variable/term (units) | Effect | SE | HR | P-value |
|---|---|---|---|---|
| **$Age^2$ ($year^2$)** | -1.4e-3 | 1.0e-4 | 0.999 | 4.6e-43 |
| **Sex (female=0, male=1)** | 2.32 | 0.247 | 10.1 | 7.0e-21 |
| **Age (year)** | 0.274 | 0.014 | 1.315 | 2.9e-84 |
| **$PGS_{CAD}$ (SD-unit)** | 1.207 | 0.181 | 3.343 | 2.5e-11 |
| **Sex*Age (year for males compared to females)** | -0.023 | 3.5e-3 | 0.978 | 1.49e-10 |
| **Sex*$PGS_{CAD}$ (SD-units for males compared to females)** | -0.423 | 0.219 | 0.655 | 0.054 |
| **Age*$PGS_{CAD}$ (SD-units per year)** | -0.013 | 2.6e-3 | 0.988 | 1.2e-6 |
| **Sex*Age*$PGS_{CAD}$ (SD-units per year for males compared to females)** | 6.0e-3 | 3.2e-3 | 1.006 | 0.062 |

SE=Standard error of the effect, HR=Hazard ratio, $PGS_{CAD}$=Coronary artery disease polygenic score, SD=Standard deviation

**Supplemental Table 11. Model statistics for additive $PGS_{CAD}$ model with ASCVD risk score (model C3)**

| Variable/term (units) | Effect | SE | HR | P-value |
|---|---|---|---|---|
| $Age^2$ ($year^2$) | -2.2e-3 | 1.3e-4 | 0.998 | 1.0e-61 |
| Sex (female=0, male=1) | 0.548 | 0.043 | 1.73 | 7.1e-38 |
| Age (year) | 0.326 | 0.016 | 1.39 | 1.0e-97 |
| ASCVD (risk/100) | 2.20 | 0.190 | 9.03 | 7.1e.31 |
| $PGS_{CAD}$ (SD-units) | 0.347 | 0.019 | 1.42 | 3.9e-77 |

SE=Standard error of the effect, HR=Hazard ratio, $PGS_{CAD}$=Coronary artery disease polygenic score, ASCVD=Atherosclerotic cardiovascular disease risk score, SD=Standard deviation

**Supplemental Table 12A-B. Cox proportional hazards model statistics for additive $PGS_{CAD}$ models for males and females separately (model S1)**

| A. Males | | | | |
|---|---|---|---|---|
| Variable/term (units) | Effect | SE | HR | P-value |
| $Age^2$ ($year^2$) | -1.4e-3 | 1.3e-4 | 0.999 | 2.4e-27 |
| Age (year) | 0.241 | 0.016 | 1.272 | 4.2e-52 |
| $PGS_{CAD}$ (SD-units) | 0.357 | 0.024 | 1.429 | 4.1e-51 |
| B. Females | | | | |
| $Age^2$ ($year^2$) | -1.3e-3 | 1.7e-4 | 0.999 | 1.90e-14 |
| Age (year) | 0.254 | 0.023 | 1.289 | 2.8e-29 |
| $PGS_{CAD}$ (SD-units) | 0.341 | 0.030 | 1.407 | 5.7e-30 |

SE=Standard error of the effect, HR=Hazard ratio, $PGS_{CAD}$=Coronary artery disease polygenic score, SD=Standard deviation

**Supplemental Table 13A-B. Cox proportional hazards model statistics for $PGS_{CAD}$ models with age interaction for males and females separately (model S2)**

| Variable/term (units) | Effect | SE | HR | P-value |
|---|---|---|---|---|
| **A. Males** | | | | |
| **Age$^2$ (year$^2$)** | -1.4e-3 | 1.3e-4 | 0.999 | 1.4e-28 |
| **Age (year)** | 0.251 | 0.016 | 1.285 | 3.9e-53 |
| **$PGS_{CAD}$ (SD-units)** | 0.780 | 0.130 | 2.182 | 1.7e-9 |
| **Age*$PGS_{CAD}$ (SD-units for one year)** | -6.5e-3 | 2.0e-3 | 0.994 | 8.9e-4 |
| **B. Females** | | | | |
| **Age$^2$ (year$^2$)** | -1.4e-3 | 1.8e-4 | 0.999 | 3.8e-16 |
| **Age (year)** | 0.275 | 0.024 | 1.317 | 2.7e-31 |
| **$PGS_{CAD}$ (SD-units)** | 1.207 | 0.184 | 3.345 | 5.7e-11 |
| **Age*$PGS_{CAD}$ (SD-units for one year)** | -0.013 | 2.6e-3 | 0.988 | 1.9e-6 |

SE=Standard error of the effect, HR=Hazard ratio, $PGS_{CAD}$=Coronary artery disease polygenic score, SD=Standard deviation

**Supplemental Table 14A-B. Model sensitivity and specificity in UK Biobank sex-stratified models.** These numbers are from models that only use samples with full 10-year follow-up (non-cases) or that get the CAD end-point during the first 10-years of follow-up (cases).

| **A. Females**<br>**Risk model** | **Incident cases with risk ⩾7.5%** | **Incident cases with risk <7.5%** | **Non-cases with risk ⩾7.5%** | **Non-cases with risk <7.5%** | **Specificity/ Selectivity** | **Sensitivity/ Recall** | **C-index [95% Confidence interval]** |
|---|---|---|---|---|---|---|---|
| **ASCVD** | 2,330 | 2,941 | 24,577 | 116,102 | 85.1% | 64.2% | - |
| **Additive $PGS_{CAD}$ model (model S1)** | 1,532 | 4,944 | 10,826 | 158,349 | 94.0% | 56.7% | 0.728 [0.722; 0.734] |
| **$PGS_{CAD}$ with age-interaction (model S2)** | 1,523 | 4,953 | 10,763 | 158,412 | 94.0% | 56.7% | 0.728 [0.722; 0.734] |
| **$PGS_{CAD}$ with age-interaction and ASCVD (model S3)** | 1,393 | 3,878 | 9,025 | 131,654 | 94.0% | 57.6% | 0.752 [0.746; 0.758] |
| **2 step: ASCVD then $PGS_{CAD}$ with age-interaction **** | 2,624 | 2,647 | 26,844 | 113,835 | 84.0% | 66.7% | - |

| **B. Males**<br>**Risk model** | **Incident cases with risk ⩾7.5%** | **Incident cases with risk <7.5%** | **Non-cases with risk ⩾7.5%** | **Non-cases with risk <7.5%** | **Specificity/ Selectivity** | **Sensitivity/ Recall** | **C-index [95% Confidence interval]** |
|---|---|---|---|---|---|---|---|
| **ASCVD** | 7,871 | 1,533 | 57,494 | 42,934 | 63.6% | 86.0% | - |
| **Additive $PGS_{CAD}$ model (model S1)** | 7,455 | 3,638 | 44,104 | 72,870 | 72.6% | 75.3% | 0.704 [0.699; 0.709] |
| **$PGS_{CAD}$ with age-interaction (model S2)** | 7,653 | 3,440 | 45,909 | 71,065 | 71.8% | 76.3% | 0.705 [0.700; 0.709] |
| **$PGS_{CAD}$ with age-interaction and ASCVD (model S3)** | 6,363 | 3,041 | 35,191 | 65,237 | 74.1% | 75.6% | 0.724 [0.719; 0.729] |
| **2 step: ASCVD then $PGS_{CAD}$ with age-interaction **** | 8,076 | 1,328 | 58,951 | 41,477 | 63.0% | 87.6% | - |

$PGS_{CAD}$=Coronary artery disease polygenic score

**Supplemental Table 15A-B. Cox proportional hazards model statistics for $PGS_{CAD}$ models with age interaction and ASCVD risk score for males and females separately (model S3)**

| Variable/term (units) | Effect | SE | HR | P-value |
|---|---|---|---|---|
| **A. Males** | | | | |
| **Age$^2$ (year$^2$)** | -2.8e-3 | 1.8e-4 | 0.997 | 4.5e-52 |
| **Age (year)** | 0.374 | 0.021 | 1.454 | 1.2e-73 |
| **$PGS_{CAD}$ (SD-units)** | 0.778 | 0.130 | 2.176 | 2.0e-9 |
| **Age*$PGS_{CAD}$ (SD-units for one year)** | -6.6e-3 | 2.0e-3 | 0.993 | 7.5e-4 |
| **ASCVD (risk/100)** | 2.965 | 0.258 | 19.39 | 1.2e-30 |
| **B. Females** | | | | |
| **Age$^2$ (year$^2$)** | -2.8e-3 | 2.4e-4 | 0.997 | 4.3e-32 |
| **Age (year)** | 0.419 | 0.030 | 1.521 | 1.9e-45 |
| **$PGS_{CAD}$ (SD-units)** | 1.210 | 0.185 | 3.352 | 5.7e-11 |
| **Age*$PGS_{CAD}$ (SD-units for one year)** | -0.013 | 2.6e-3 | 0.987 | 1.2e-6 |
| **ASCVD (risk/100)** | 3.081 | 0.314 | 21.77 | 1.1e-22 |

SE=Standard error of the effect, HR=Hazard ratio, $PGS_{CAD}$=Coronary artery disease polygenic score, SD=Standard deviation, ASCVD=Atherosclerotic cardiovascular disease risk score

#### Supplemental Figures

**Supplemental Figure 1A-B.** Distribution of the Coronary artery disease polygenic score ($PGS_{CAD}$). Panel A shows the distribution of the raw $PGS_{CAD}$ and panel B distribution after PC-adjustment. N=Number of samples.

A. B.

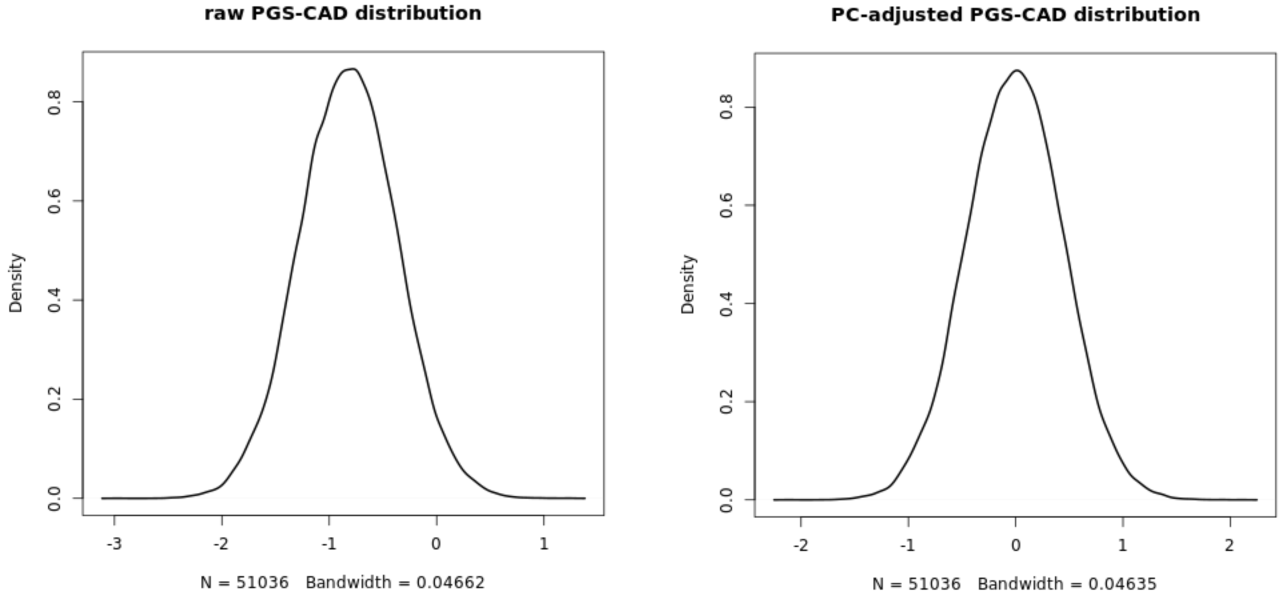


**Supplemental Figure 2.** Calibration of the ASCVD risk score/Pooled Cohort Equation in HUNT2 dataset. Panel A shows the calibration of the full risk distribution and panel B within risks < 20%. Each dot in the figure represents 5% of the predicted risk distribution.

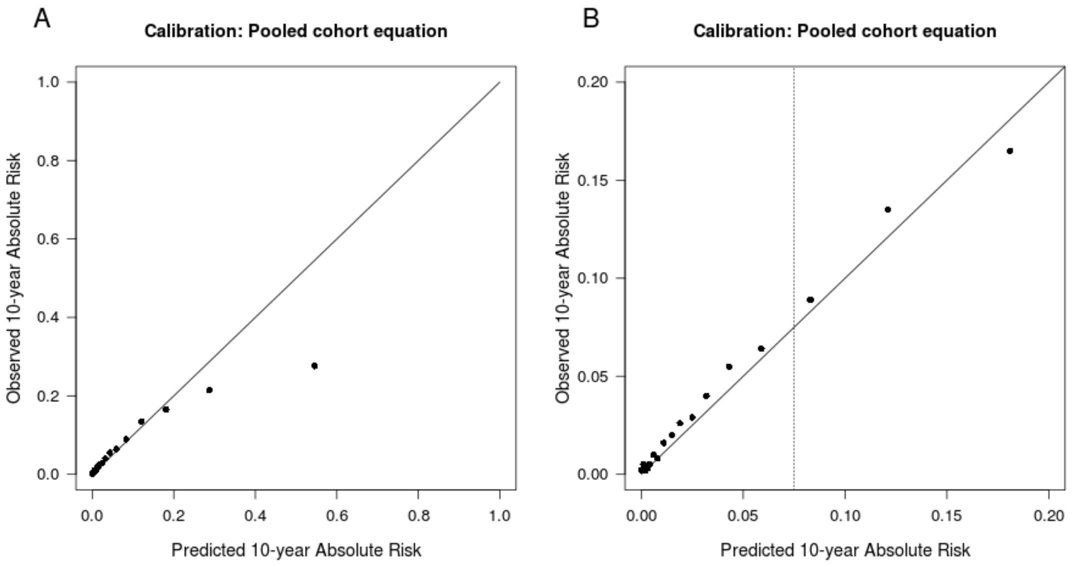


**Supplemental Figure 3. Raw $PGS_{CAD}$ by age and sex zoomed in Y-axis**

Illustration of the selection bias in the cohort baseline showing generalized additive model (GAM) curves for $PGS_{CAD}$ by age for males and females separately overlaid hexagonal bins of sample counts in the UKBB replication dataset. Panel A is for the analysis dataset used in the Cox models and panel B when prevalent cases are included.

**A.**

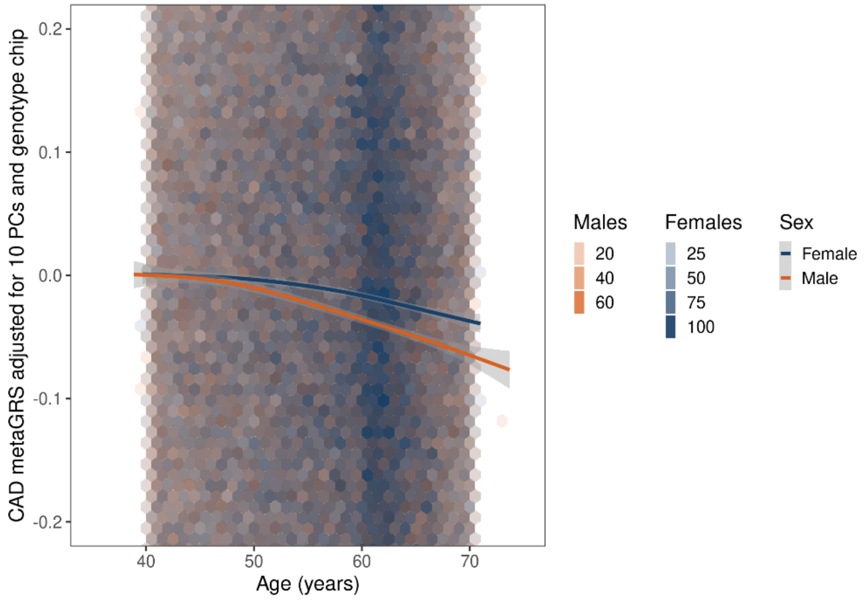


**B.**

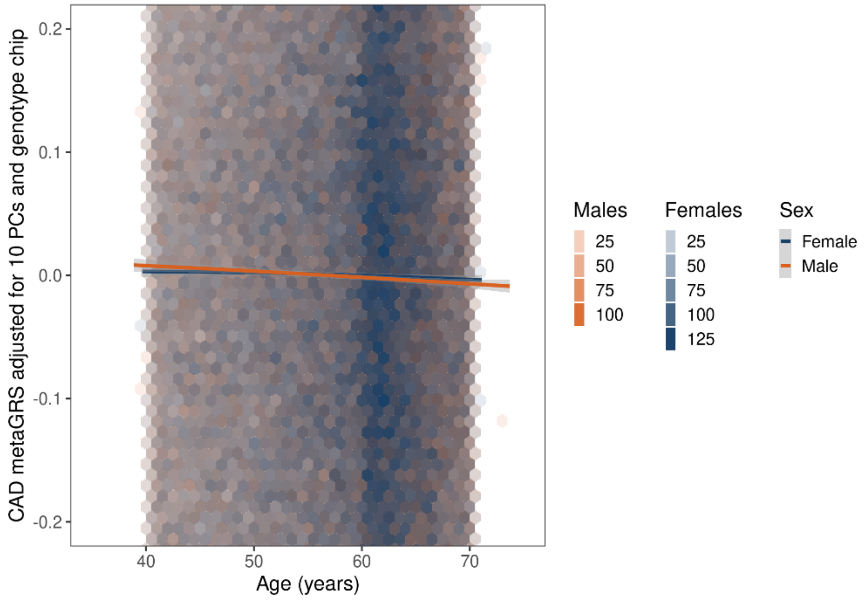


**Supplemental Figure 4. The effect of threshold on the sensitivity and specificity of model S1 (Additive PGS$_{CAD}$ model).** The panel A shows sensitivity, panel B specificity and panel C the proportion of individuals in the high-risk group for males (in blue) and females (in red) separately.

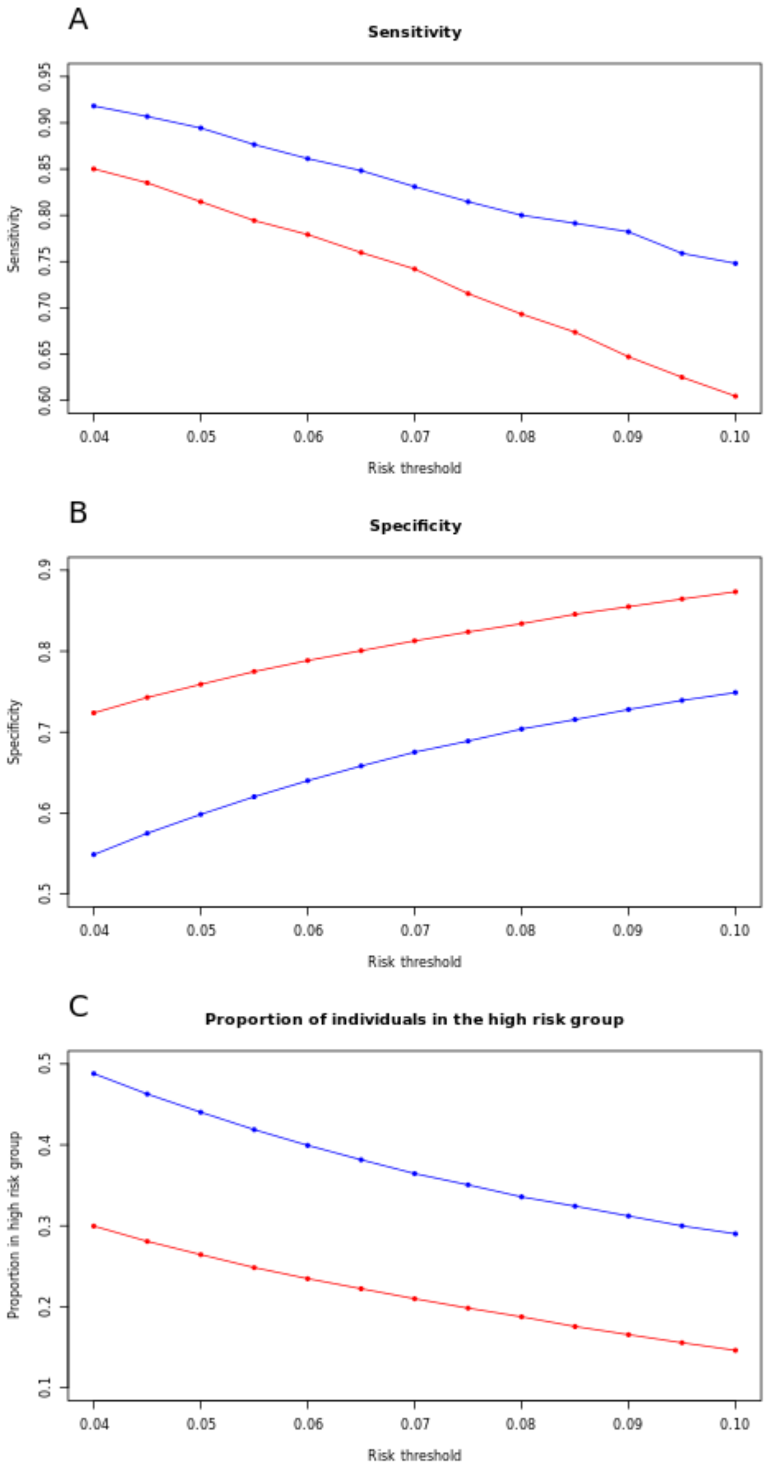
A
Sensitivity
Sensitivity
0.60
0.65
0.70
0.75
0.80
0.85
0.90
0.95
0.04
0.05
0.06
0.07
0.08
0.09
0.10
Risk threshold
B
Specificity
Specificity
0.5
0.6
0.7
0.8
0.9
0.04
0.05
0.06
0.07
0.08
0.09
0.10
Risk threshold
C
Proportion of individuals in the high risk group
Proportion in high risk group
0.1
0.2
0.3
0.4
0.5
0.04
0.05
0.06
0.07
0.08
0.09
0.10
Risk threshold

**Supplemental Figure 5. The effect of threshold on the sensitivity and specificity of model S2 ($PGS_{CAD}$ with age-interaction).** The panel A shows sensitivity, panel B specificity and panel C the proportion of individuals in the high-risk group for males (in blue) and females (in red) separately.

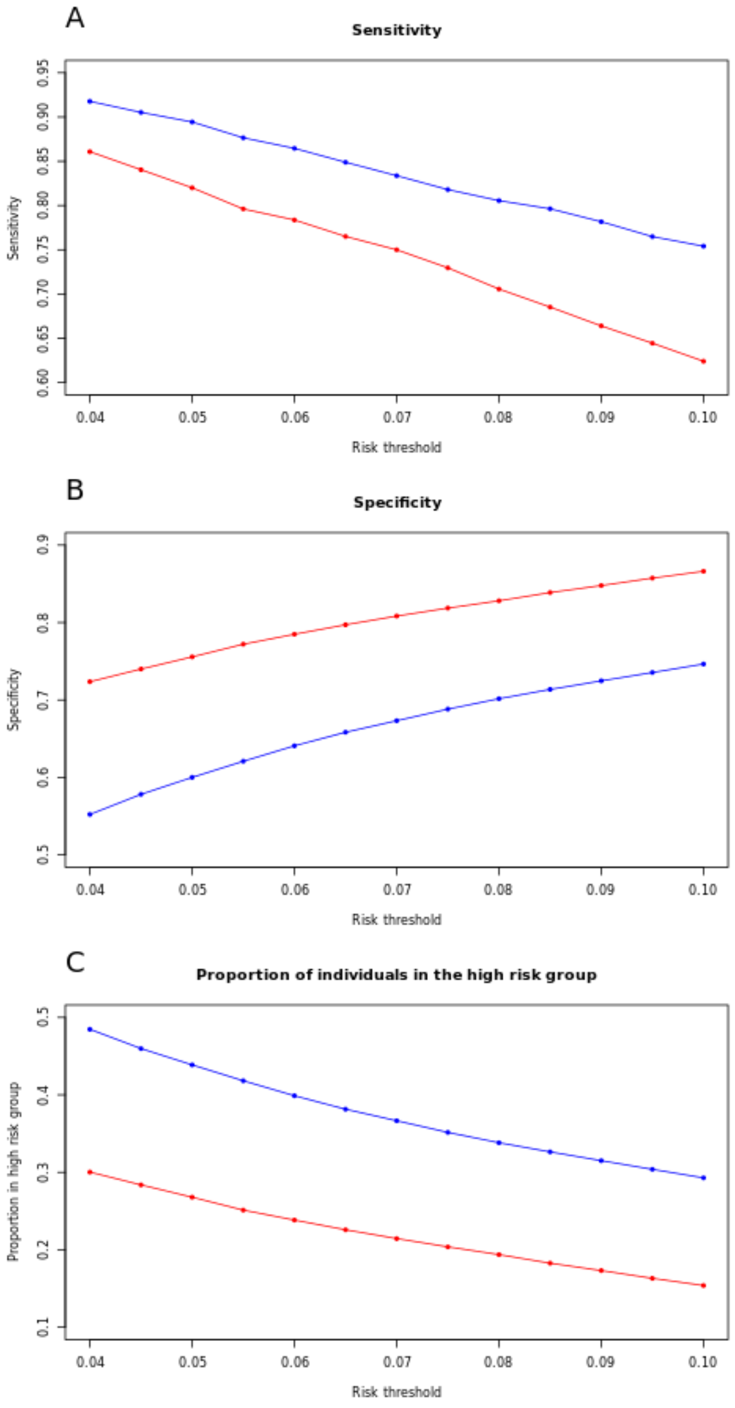

**Supplemental Figure 6. The effect of threshold on the sensitivity and specificity of model S3 ($PGS_{CAD}$ with age-interaction and ASCVD risk score).** The panel A shows sensitivity, panel B specificity and panel C the proportion of individuals in the high-risk group for males (in blue) and females (in red) separately.

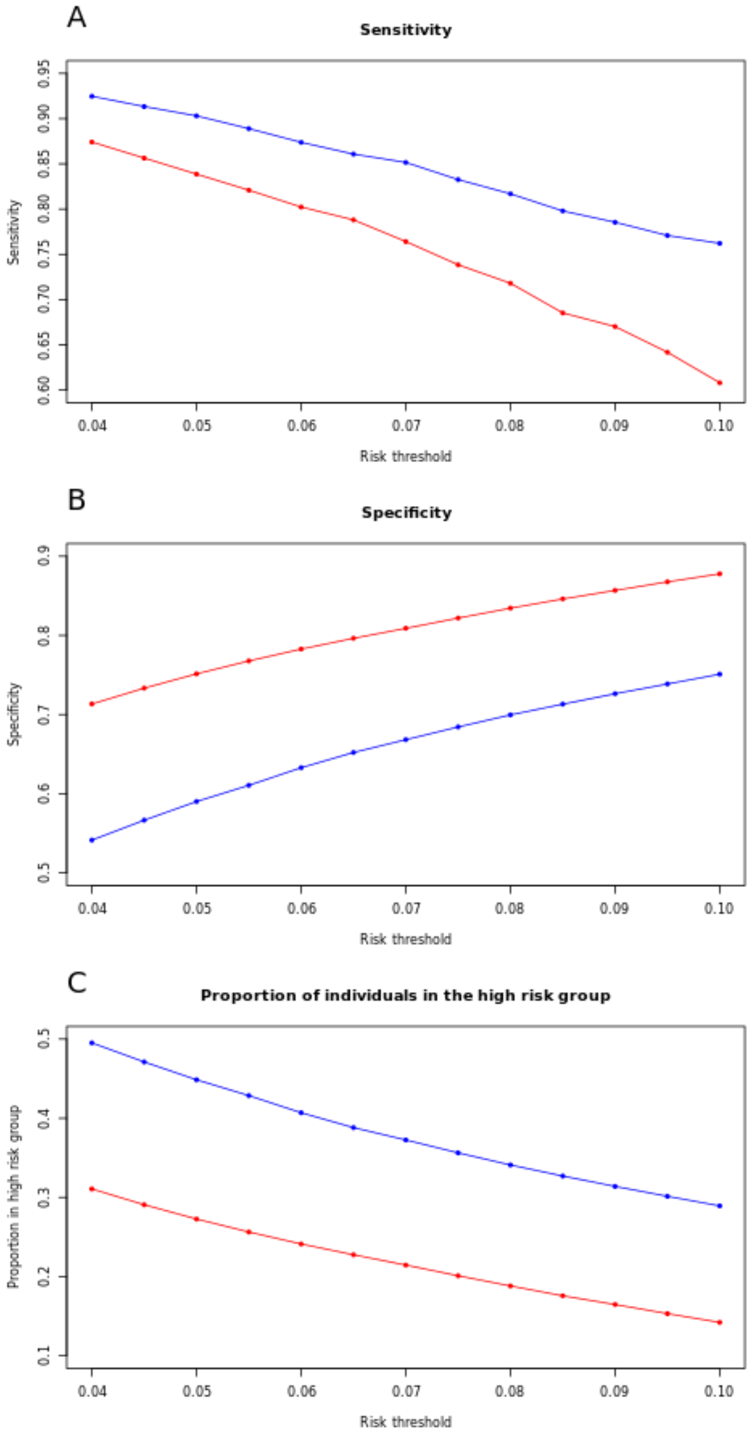

**Supplemental Figure 7. The effect of threshold on the sensitivity and specificity of ASCVD risk score.** The panel A shows sensitivity, panel B specificity and panel C the proportion of individuals in the high-risk group for males (in blue) and females (in red) separately.

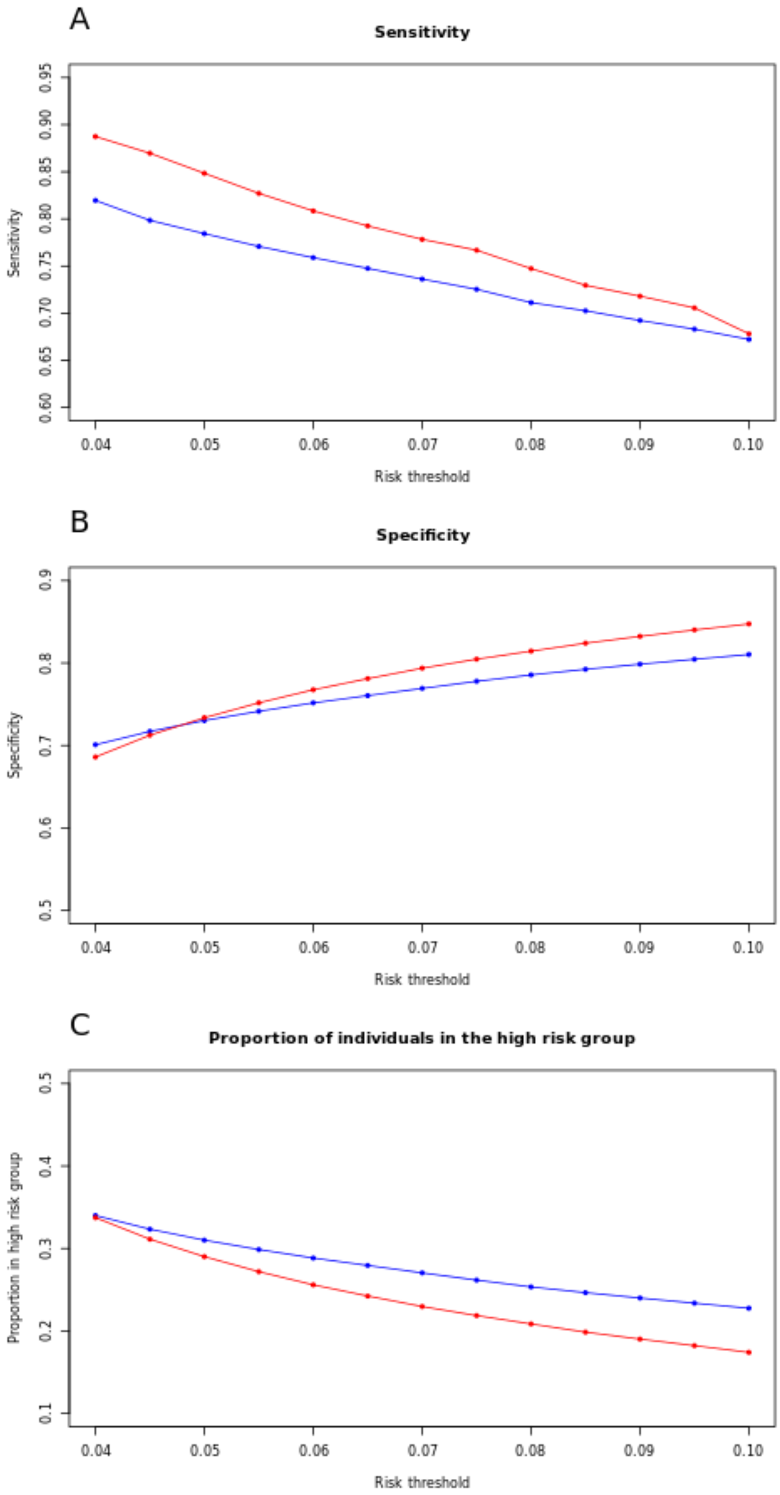

**Supplemental Figure 8. Calibration of the ASCVD risk/Pooled Cohort Equation for males and females separately**. The figures are restricted to show risks < 20% and the risk threshold used in the study (7.5%) has been highlighted with a dashed line. Panel A shows the calibration for females and panel B for males.

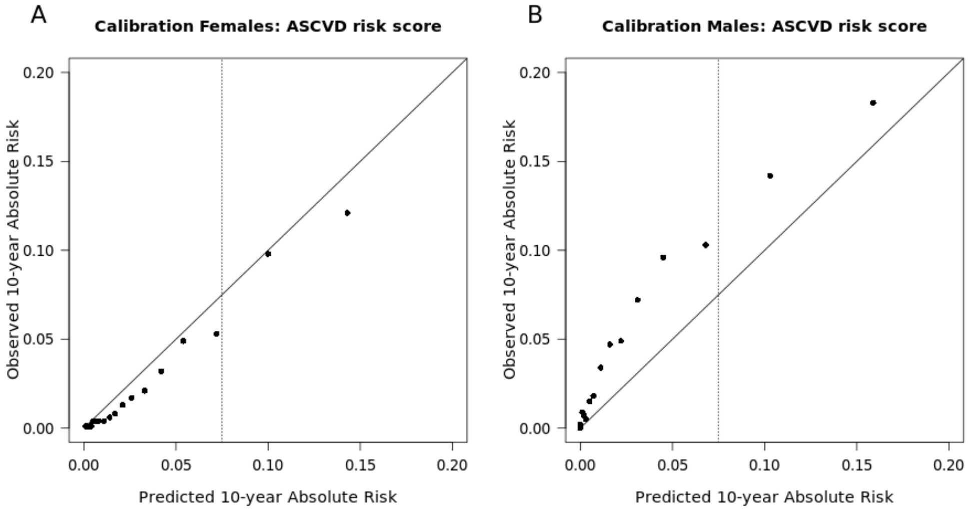


**Supplemental Figure 9. Calibration of the model S3 ($PGS_{CAD}$ with age-interaction and ASCVD risk score) for males and females separately**. The figures are restricted to show risks < 20% and the risk threshold used in the study (7.5%) has been highlighted with a dashed line. Panel A shows the calibration for females and panel B for males.

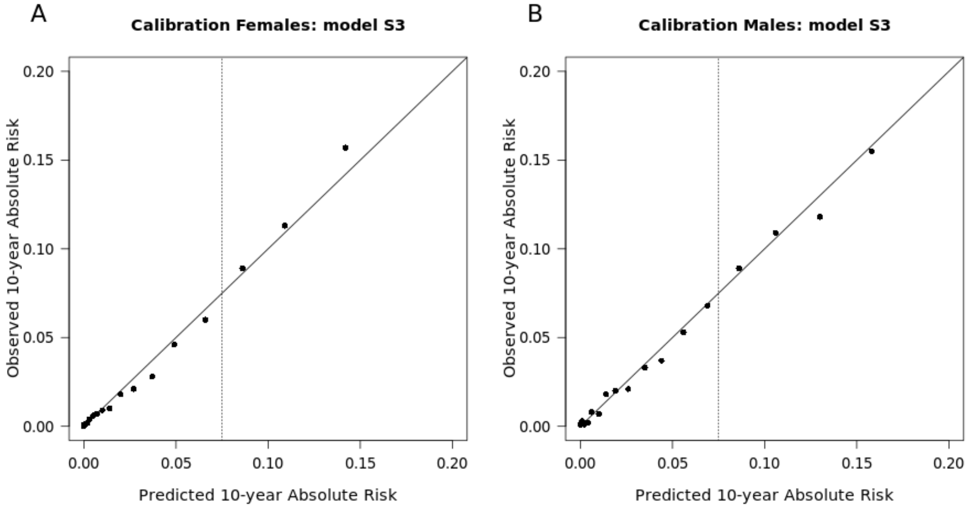
